# How is the COVID-19 pandemic impacting our life, mental health, and well-being? Design and preliminary findings of the pan-Canadian longitudinal COHESION Study

**DOI:** 10.1101/2022.05.26.22275645

**Authors:** Stephan Gabet, Benoit Thierry, Rania Wasfi, Margaret De Groh, Guido Simonelli, Catherine Hudon, Lily Lessard, Ève Dubé, Bouchra Nasri, Yan Kestens, Grégory Moullec

## Abstract

With the advent of the COVID-19 pandemic, in-person social interactions and opportunities for accessing resources that sustain health and well-being have drastically reduced. We therefore designed the pan-Canadian population-based prospective *COVID-19: HEalth and Social Inequities across Neighbourhoods* (COHESION) cohort to provide deeper understanding of how the COVID-19 pandemic context affects mental health and well-being, key determinants of health, and health inequities.

This paper presents the design of the two-phase COHESION Study, and descriptive results from the first phase conducted between May 2020 and September 2021. During that period, the COHESION research platform collected monthly data linked to COVID-19 such as infection and vaccination status, perceptions and attitudes regarding pandemic-related measures, and information on participants’ physical and mental health, well-being, sleep, loneliness, resilience, substances use, living conditions, social interactions, activities, and mobility.

The 1,268 people enrolled in the Phase 1 COHESION Study are for the most part from Ontario (47%) and Quebec (33%), aged 48 ± 16 years [mean ± standard deviation (SD)], and mainly women (78%), White (85%), with a university degree (63%), and living in large urban centers (70%). According to the 298 ± 68 (mean ± SD) prospective questionnaires completed each month in average, the first year of follow-up reveals significant temporal variations in standardized indexes of well-being, loneliness, anxiety, depression, and psychological distress.

The COHESION Study will allow identifying trajectories of mental health and well-being while investigating their determinants and how these may vary by subgroup, over time, and across different provinces in Canada, in the unique context of the COVID-19 pandemic.

## 1. Introduction

On January 10, 2020, WHO announced the identification of a new strain of coronavirus, the severe acute respiratory syndrome coronavirus 2 (SARS-CoV-2), causing a mild to severe respiratory illness – which may progress to pneumonia and respiratory failure – named coronavirus disease 2019 (COVID-19) [1]. The first positive case of COVID-19 in Canada was reported on January 25, 2020 and a state of health emergency was declared across the country between March 13 and March 22, 2020, depending on the province [2,3]. Mid-August 2021, there were just over 1.4 million cases in Canada and more than 26,700 deaths [2].

Public health measures to control transmission have brought vast sectors of economic activity to a halt, leading to massive unemployment and reductions in income, while reducing people’s daily movements and opportunities for in-person social interactions [4]. This has affected people’s opportunities and access to resources that sustain mental health and well-being. The COVID-19 conditions reinforce the role of various social and environmental health determinants, with differential impacts on the mental health and well-being of populations, depending on age [5], gender [6], housing conditions [7], education [8], job type [9], income [10], or ethnic background [11–13] and, more generally, social and environmental contexts [14,15].

COVID-19 conditions including lockdown, curfew and physical distancing measures reduce social contacts, increase social isolation and feelings of loneliness, and decrease levels of social support; these dimensions directly influence well-being and mental health [16,17]. Canadian data shows pandemic-related increases in social isolation, domestic violence and anxiety [18], with certain groups such as teens, older adults, women and racialized communities particularly at-risk [19]. Daily mobility and related physical activity are reduced through confinement measures and reduced activity spaces have been linked to depressive symptoms [20,21] and sleep troubles [22]. Walkable environments and access to green space are key environmental conditions linked to positive health outcomes including physical activity and well-being [23]. With shrinking activity spaces and policy constraints curbing daily mobility, the role of residential living conditions is further amplified.

Priority populations often bear the burden of poor social and environmental living conditions and have been shown to be disproportionately affected by COVID-19 [24,25]. Overcrowding or living in inadequate dwellings are important determinants of mental health and well-being; moreover, prolonged exposure to home environments during lockdown conditions further exacerbate these impacts [26,27]. Economic hardship, which is linked to income reduction and job instability, has heavily contributed to the mental health burden of Canadians, and is also related to housing instability and food insecurity [28,29]. In turn, detrimental health behavior such as alcohol or other substance use are increasing and are linked to poorer mental health outcomes [30].

In total, there is an urgent need to better understand, in particular, how the unintended long-term consequences of COVID-19 pandemic and mitigation measures, contextual conditions (e.g., housing conditions, neighborhood characteristics), and behavior (e.g., mobility, social interaction, sleep) are linked to mental health and well-being trajectories. Furthermore, impacts are likely to vary between population groups (for instance, according to gender, age, racialized communities, or deprivation level).

## 2. Two-phase COHESION Study objectives

We developed the pan-Canadian *COVID-19: HEalth and Social Inequities across Neighbourhoods* (COHESION) Study to better understand how the COVID-19 pandemic affects health, key determinants of health, and health inequities, with a focus on mental health and well-being. This study will provide longitudinal evidence of how these change over time and across different provinces in Canada.

In the context of the COVID-19 pandemic, the COHESION Study aims (Fig 1):

- evaluating the direct impact of physical and social neighborhood characteristics on mental health and well-being trajectories, while controlling for individual-level health behaviors and socio-demographics;
- evaluating if and how physical and social neighborhood characteristics may modify the associations between individual-level predictors and mental health and well-being trajectories.

**Figure 1.**
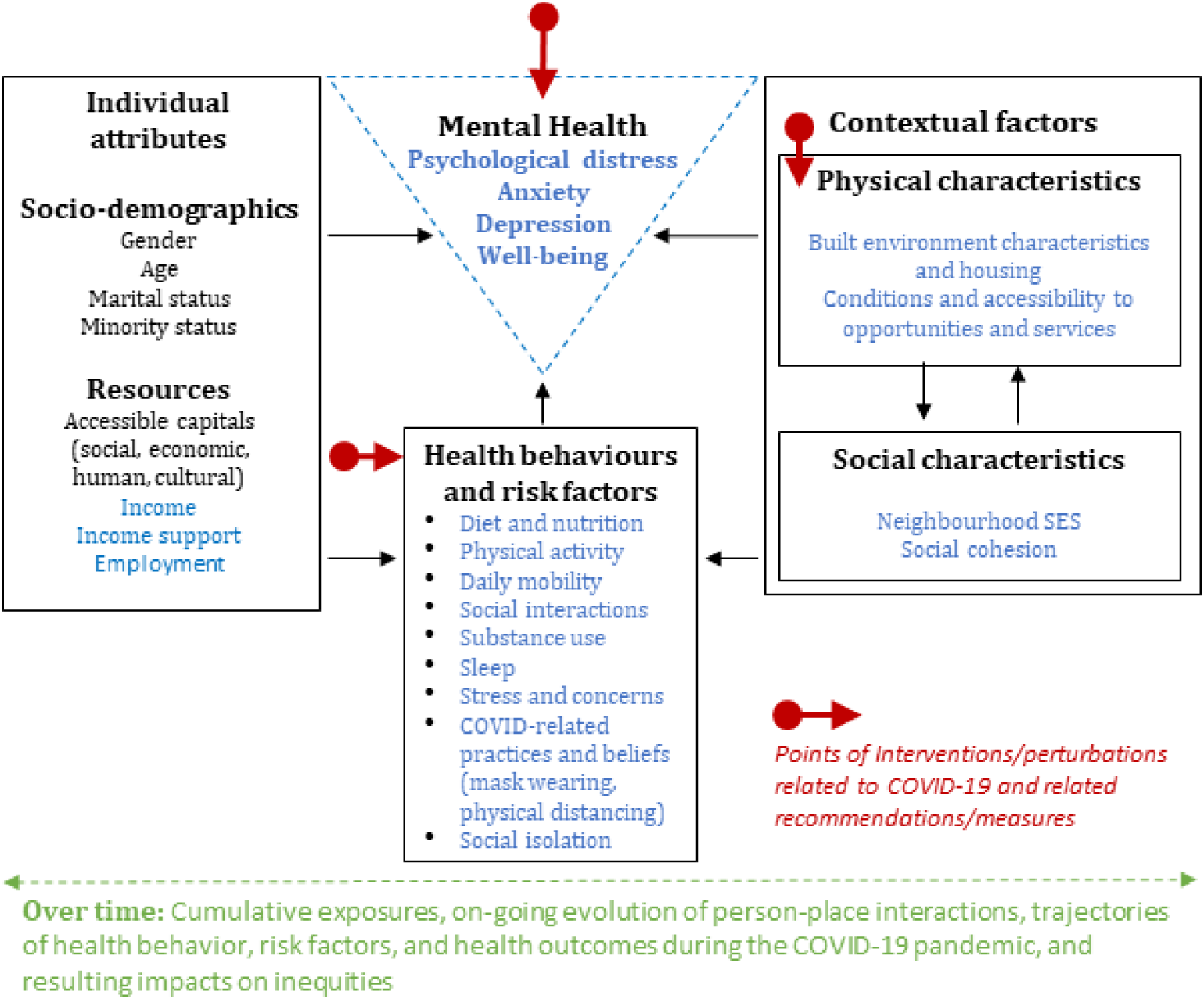
Conceptual model of the COHESION Study. Adapted from Wasfi & Kestens (2021) [65].

This paper presents the design and methods used in the two phases of the COHESION study. It also reports descriptive statistics from the longitudinal follow-up of the first phase of the study and lessons learned from its implementation.

## 3. Two-phase COHESION Study design

### 3.1. Study design and recruitment

The two-phase COHESION Study is a pan-Canadian population-based prospective cohort study. COHESION Phase 1 was conducted between May 2020 and September 2021. COHESION Phase 2 will be launched in May 2022. All Phase 1 participants will be invited to participate in Phase 2, and additional recruitments (n = 10,000) will be done.

After consenting to participate to the study, participants are invited to fill a short eligibility questionnaire. Inclusion criteria are being aged 15 years or above, currently residing in Canada, and reading or speaking English or French. Eligible participants are then invited to complete a baseline questionnaire.

The recruitment for COHESION Phase 1 was launched on May 11 2020. We used a combination of methods that have proven successful from previous experience in recruiting population-based research project samples [31], including media communication (e.g., newspaper articles, radio interviews), social media promotion (e.g., Facebook, Twitter, Instagram, and LinkedIn), and outreach through partners’ local networks (e.g., local health authorities that relayed the study on their website or through their newsletters).

For COHESION Phase 2, we will further use quota sampling at the health region level, based on 2016’s Census data (i.e., age composition, gender, income, educational attainment, and ethnic background). We engaged Potloc Inc., a tech-enabled consumer research company that conducts survey sampling through social networks (Facebook, Twitter, Instagram, LinkedIn), for geo-targeting of respondents based on the sampling quotas. Potloc Inc.’s algorithm will push sociodemographic and geographically targeted online ads to our study until local quotas are attained (targets are monitored daily) and that 10,000 participants have thereby been enrolled.

Enrollment and participation in the two-phase COHESION Study are voluntary, and a raffle of three prizes ($100 gift cards) is drawn every month for active participants for the duration of the study.

### 3.2. Data collection

#### 3.2.1. Involvement options

Two levels of involvement are proposed. Participants can opt for: i) participating in the online self-administered questionnaires only (baseline + invitation to follow up questionnaires); or ii) participating in the questionnaires and downloading a mobile phone application to provide additional active and passive data (cf. 3.2.5).

#### 3.2.2. Baseline questionnaires

Through the COHESION Study Phase 1 baseline questionnaire (35-45 min), participants report on: their sociodemographic characteristics (e.g., gender, ethnic or cultural background, education level, employment status, income, relationship status, and household composition); housing condition (e.g., house tenure, dwelling type and size, outdoor space) and satisfaction; physical (e.g., chronic disease diagnosis) and mental health conditions (e.g., anxiety and depression), and substance use (alcohol, tobacco, vape, and cannabis); COVID-19 infection status, vulnerability towards COVID-19, perception of- and compliance with COVID-19 mitigation measures, and pre-pandemic employment, income, substances use, loneliness, resilience, and social interactions. Additionally, a map-based questionnaire collects data on activity locations and social contacts (cf. 3.2.4). For parents, a supplementary module on mental health and perceptions of COVID-19 mitigation measures concerning adolescents and children who live in the household is administered (participants answer on behalf of children in their household).

The COHESION Study Phase 2 baseline questionnaire will consist in one core (10 min) and two optional complementary sections (15 min each). The core baseline questionnaire includes: sociodemographic characteristics and key housing conditions; COVID-19 vaccination status, perceptions of COVID-19 pandemic and related mitigation measures; standardized modules assessing general health (first item of the 12-item Short-Form Health Survey) [32], well-being (5-item World Health Organization Well-Being Index, WHO-5 Index) [33], sleep credit (Pittsburgh Sleep Quality Index, PSQI, 4 items only) [34], and psychological distress (Psychological Distress Scale, Kessler-6) [35]; and an outdoor mobility and social interactions module. Optional complementary baseline sections include the map-based questionnaire on activity locations and social contacts (cf. 3.2.4) and additional in-depth questions on relevant themes: additional items retrieved from the Phase 1 baseline questionnaire (it means all items not already included in the Phase 2 core baseline questionnaire); standardized modules on loneliness (University of California Los Angeles, UCLA, 3-item loneliness score) [36], anxiety symptoms (7-item Generalized Anxiety Disorder, GAD-7) [37], depression symptoms (9-item Patient Health Questionnaire, PHQ-9) [38], resilience (6-item Brief Resilience Scale) [39], sense of belonging (Canada Community Health Survey, CCHS) [40], and physical activity (Godin Leisure-Time Exercise) [41]; items for assessing conspiracy beliefs.

Slightly shorten versions of the Phase 2 baseline questionnaires (i.e., excluding questions about birth, ethnics, etc.) have been provided for participants coming from Phase 1 and wishing going on Phase 2.

#### 3.2.3. Follow-up questionnaires

Follow-up questionnaires of the COHESION Study are short questionnaires (15 min). For Phase 1, they were first offered biweekly until August 2020, then monthly. COVID-19-related topics cover infection and/or vaccination status, perceived vulnerability, perception of and compliance with mitigation measures, position regarding vaccination, and conspiracy beliefs. Health-related questions focus on general health (SF-12, first item) [32], well-being (WHO-5 Index) [33], sleep credit (PSQI, 4 items only) [34], loneliness (UCLA 3-item loneliness score) [36], anxiety symptoms (GAD-7) [37], depression symptoms (PHQ-9) [38], psychological distress (Kessler-6) [35], and sense of belonging (CCHS) [40] (Table 1). Employment status, household income, and substance use are also documented. Additionally, participants are asked at each follow-up about any changes concerning their place of residence and housing conditions. Supplementary module on mental health and perception of COVID-19 mitigation measures concerning children or adolescents living at home is administered when applicable.

**Table 1.** Main information collected in the framework of the COHESION Study Phase 1 prospective follow-up (June 2020 to July 2021).

|  |  |  | Survey (year/week) |  |  |  |  |  |  |  |  |  |  |  |  |  |  |  |  |
| --- | --- | --- | --- | --- | --- | --- | --- | --- | --- | --- | --- | --- | --- | --- | --- | --- | --- | --- | --- |
| Theme | Assessment tool | Assessment period | 20/26 | 20/30 | 20/32 | 20/34 | 20/36 | 20/38 | 20/40 | 20/44 | 20/48 | 20/52 | 21/03 | 21/07 | 21/11 | 21/15 | 21/19 | 21/23 | 21/27 |
| COVID-19 |  |  |  |  |  |  |  |  |  |  |  |  |  |  |  |  |  |  |  |
| COVID-19 infection status |  | At the questionnaire administration point | X | X | X | X | X | X | X | X | X | X | X | X | X | X | X | X | X |
| COVID-19-pandemic-related perceptions |  | At the questionnaire administration point | X | X | X | X | X | X | X | X | X | X | X | X | X | X | X | X | X |
| Anti-COVID-19 measures compliance |  | At the questionnaire administration point |  |  |  | X | X | X | X | X | X | X | X | X | X | X | X | X | X |
| Position towards anti-COVID-19 vaccine |  | At the questionnaire administration point |  |  |  |  |  |  | X |  |  | X |  |  |  | X | X | X | X |
| Anti-COVID-19 vaccination status |  | At the questionnaire administration point |  |  |  |  |  |  |  |  |  |  |  |  | X | X | X | X | X |
| Health / Well-being |  |  |  |  |  |  |  |  |  |  |  |  |  |  |  |  |  |  |  |
| General health | SF-12 (first item only) | During the past 2 weeks | X | X | X | X | X | X | X | X | X | X | X | X | X | X | X | X | X |
| Well-being | WHO-5 Index | During the past 2 weeks |  |  |  |  |  |  |  |  |  | X |  |  |  | X | X | X | X |
| Loneliness | UCLA 3-item loneliness score | During the past 2 weeks |  |  |  |  | X | X | X | X | X | X | X | X | X | X | X | X | X |
| Anxiety symptoms | 7-item General Anxiety Disorder (GAD-7) | During the past 2 weeks | X | X |  |  |  | X |  |  |  | X |  |  |  | X | X | X | X |
| Depression symptoms | 9-item Patient Health Questionnaire (PHQ-9) | During the past 2 weeks | X | X |  |  |  | X |  |  |  | X |  |  |  | X | X | X | X |
| Psychological distress symptoms | 6-item Psychological Distress Scale (Kessler-6) | During the past 2 weeks |  |  | X | X | X | X | X | X | X | X | X | X | X |  |  |  |  |
| Behaviors |  |  |  |  |  |  |  |  |  |  |  |  |  |  |  |  |  |  |  |
| Outdoor mobility | VERITAS | During the past 7 days | X | X | X | X | X | X | X | X | X | X | X | X | X | X | X | X | X |
| Social interactions | VERITAS | During the past 7 days | X | X | X | X | X | X | X | X | X | X | X | X | X | X | X | X | X |
| Sleep credit | Pittsburgh Sleep Quality Index | During the past month | X | X |  |  |  | X |  |  |  | X |  | X |  | X | X | X | X |
| Substance use <sup>a</sup> | Frequency questionnaire | During the past 2 weeks |  |  | X |  | X |  | X | X |  | X |  | X |  | X | X | X | X |

<sup>a</sup> Four items: alcohol, cigarettes, vape, and cannabis.Follow-up questionnaire waves are named according to their week and year of release (for instance, “20/26” for the follow-up questionnaire proposed to participants in the 26<sup>th</sup> week of 2020).

For Phase 2, follow-up questionnaires will be split in a core (‘light’) and an optional complementary (‘complete’) sections, and will be offered every two months. It will include the same questions as for Phase 1, adding a standardized module for assessing physical activity (Godin Leisure-Time Exercise Questionnaire) [41].

Questions can vary between follow-ups, depending on the situation and priorities raised by the research team and our public health partners. Thus, these additional themes can focus on health insecurity (i.e., access to health care, prescriptions and medicine) [42], sleep troubles (PSQI) [34], food insecurity (10-item Health Canada Household Food Security Module) [43], and children’s difficulties (if any) [44].

#### 3.2.4. Use of VERITAS-Social to collect daily mobility and social interaction data

One of the specificities of COHESION is that it integrates, in baseline and follow-up questionnaires of the two phases, the *Visualization and Evaluation of Route Itineraries, Travel destinations, Activity spaces and Social interactions* (VERITAS-Social) questionnaire. For COHESION, it was adapted to locate a possible list of up to 20 activities carried out during the past seven days (Fig 2). As previously described, VERITAS-Social is an interactive map-based questionnaire that jointly collects an individual’s social network and activity locations [44]; it is an adaptation of the VERITAS tool, an interactive questionnaire for geo-locating places, and related information of interest (e.g., frequency of visit, transportation modes used) [45]. It uses a Google Map module to facilitate the location of activity places. The social module asks if an activity location is generally visited alone or with someone else; participants can identify one or more individuals, or a group of people (*see* S1a-b Figs). In other words, it is a name generator that identifies network members based on their co-presence at reported destinations [46]. Data on network members include age, gender, type of relationship (e.g., friend, acquaintance), frequency of interactions, and duration of the relationship; for groups, data includes the number of people in the group, and the duration of the relationship.

**Figure 2.**
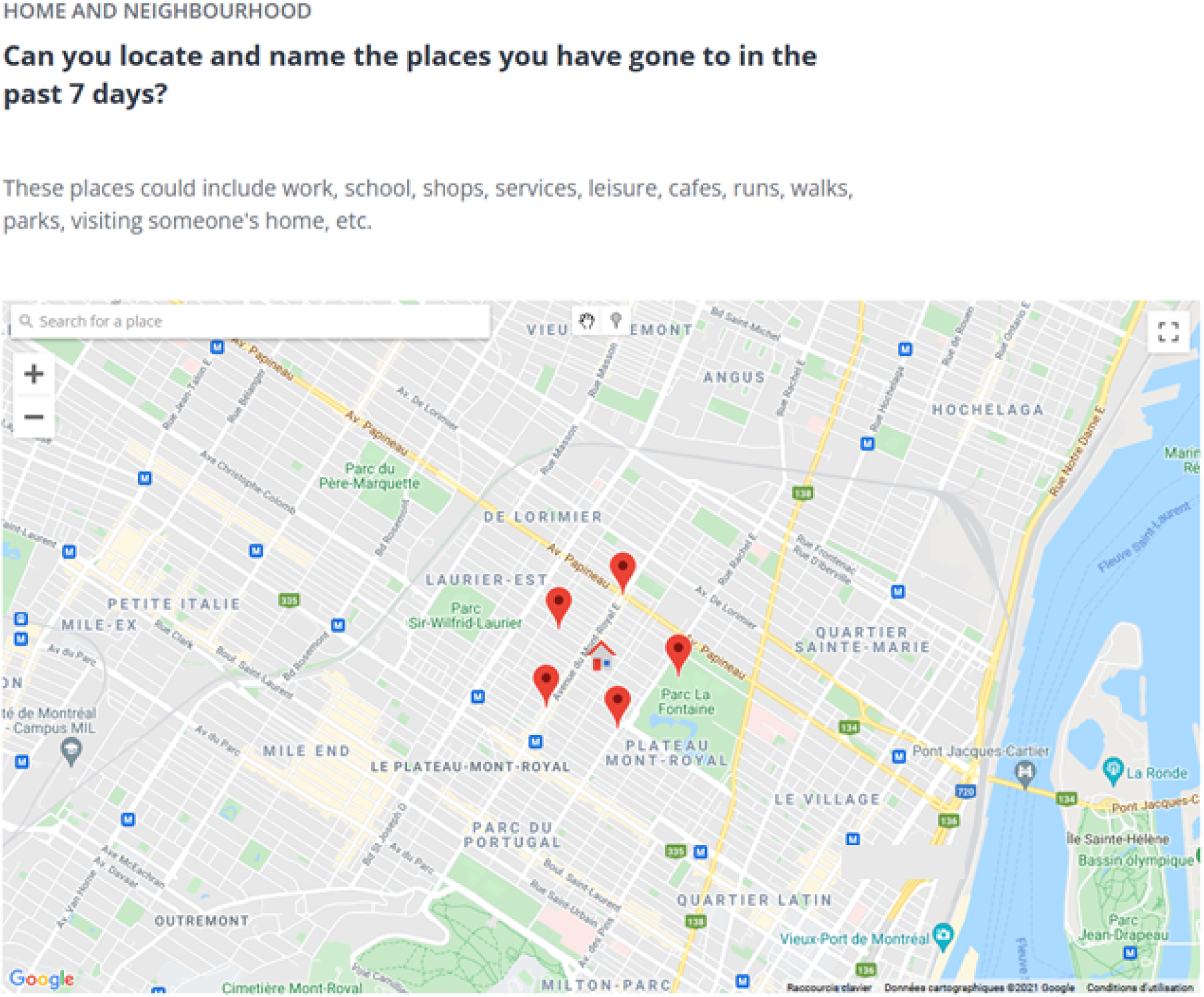
Collecting data on living and activity places with VERITAS-Social. Screenshot for a fictional participant; the house icon locates the participant’s residential address and the pin icons symbolize the visited places located by the participant.

Once all activities, corresponding locations, and all people with whom these activities are carried out have been reported, participants are asked to identify members of their social network from whom they receive support or with whom they enjoy spending time with, including members who may not have been reported among the people seen at usual activity venues. Finally, participants were able to indicate interpersonal relationships between network members (i.e., who knows whom), and whether some specific individuals belonged to documented groups.

#### 3.2.5. Optional mobile application

Participants enrolling in the mobile phone arm of the study were invited to download Ethica Data mobile application (cf. 3.2.1), used successfully in previous research projects [47]. Once installed and launched, the application passively monitors location and mobility (e.g., stationary, in vehicle, walking, or biking) using GPS data (1 minute of data collected every 5 minutes), physical activity (step counter), and social contacts (other smartphones), derived from listings of other Bluetooth discoverable devices in the participant’s surroundings [48]. The app further prompts participants to complete Ecological Momentary Assessment (EMA) questionnaires three times a day for one week every month. EMA questionnaires include short self-reports (<1 min) on well-being, sleep, mood (Short Mood Scale, 6 items) [49], and social interactions (*see* S2 Fig).

#### 3.2.6. Environmental data

Because some of the data collected is spatial (e.g. place of residence, VERITAS-Social locations, mobile app GPS data), we are able to add area-level environmental data to our dataset. This allows to explore links between individual-level measures and social and environmental contextual conditions. Numerous environmental datasets covering Canada are available through the *Canadian Urban Environmental Health Research Consortium* (CANUE).

Contextual variables of interest include measures of neighborhood deprivation, such as the Material and Social Deprivation Indices (MSDI; data 2016) available at the Canadian Census dissemination area level (i.e., the smallest Canadian standard geographic area, with a population of 400 to 700 persons) [50]. These two composite indexes combine Census data on educational attainment, employment ratio, average income, and household composition.

The urbanization degree is measured using *Statistics Canada* classification (at the four-digit code area level; data 2016), based on the number of inhabitants in population centers [51]. “Small”, “medium” and “large” urban population centers correspond to areas embracing between 1,000 and 29,999, between 30,000 and 99,999, and 100,000 and more inhabitants, respectively, while “rural area” is a residual value gathering all areas located outside population centers.

Greenness is evaluated using the growing season Normalized Difference Vegetation Index (NDVI) (at the six-digit code area level; data 2019), based on Landsat 8 satellite data [52,53].

Area-level walkability is measured using the *Canadian Active Living Environments* (CAN-ALE) database [54].

Additional localized and temporalized data on COVID-19 pandemic conditions, including lockdowns, curfews, vaccination plan, number of cases, and emergence of variants is also available through the *Canadian Institute for Health Information* (CIHI) compilation [55].

### 3.3. Ethics and data management plan

Before completing the eligibility questionnaire, potential participants are first invited to read the consent form and provide an electronic written consent to participate in the study. In Phase 2, a second consent will be required from participant wishing to register for the prospective follow-ups after having completed baseline questionnaire. Phase 1 COHESION was approved by the ethics board of the *Centre de Recherche du Centre Hospitalier de l’Université de Montréal* (CRCHUM; MP-02-2021-8924) and by the *Public Health Agency of Canada* (PHAC) ethical review board (REB 2020-016P). Phase 2 COHESION was approved by both the ethics board of the *Centre Intégré Universitaire de Santé et de Services Sociaux du Nord-de-l’Île-de-Montréal* (CIUSSS-NIM ; 2022-2327) and PHAC (REB 2020-016P).

For data security and privacy proposal, all data are hosted on *Compute Canada* servers with secondary backups on hospital-grade internal servers. All data has been stripped from any personally identifying information, with only the principal investigator being able to track the records back to one participant. Researchers can access anonymized individual level records by connecting directly through a Secure Shell (SSH) to the database hosted on *Compute Canada* (a SSH allows remote machines accessing data in a secure way since the connection is encrypted). They can also access aggregated data for analysis through a secure online platform hosted by Tableau [56]. A document listing all the data management policies governing data access and storage has been submitted to the ethic board for approval.

## 4. COHESION Study Phase 1 first results

### 4.1. Recruitment and participants

Among the 2,557 people who completed the COHESION Phase 1 eligibility questionnaire, 2,346 (92%) met the selection criteria (Fig 3). Among these ones, 1,268 (54%) completed the baseline questionnaire and have been enrolled in the COHESION Study, and among the latter, 557 (44%) opted for the Ethica mobile application in addition to the online self-administered questionnaires. Although recruitment was continuously open during the Phase 1 timespan, the major part of participants joined the study during the first months following the study launch: 832 (66%) recruitments after three months of follow-up, 1,136 (90%) after six months (Fig 4). COHESION Phase 1 participants are distributed all across Canada; they live mainly in Ontario (597, 47%) and Quebec (417, 33%) (Fig 5), and 910 (72%) declared English as their favorite language. They are in average 48 ± 16 (mean ± standard deviation, SD) and are mainly women (78%), White (85%), born in Canada (85%), with university or post-graduate level degree (63%), and in a relationship (67%) (Table 2a). The most part is owner (62%) and lives in a house (66%), with their partner or family (74%), without children at home (72%), with private outside space access (97%), with pets (56%), and in large urban centers (i.e., in areas including more than 100,000 inhabitants); though participants show wide contrasts in terms of home surroundings’ greenness and neighborhood’s material and social deprivation (Table 2b). Participants are, by decreasing order, employed (58%), retired (19%), unemployed (14%), on leave or disabled (5%), or students (2%); they are mostly satisfied about their household annual income (77%) (Table 2a). Concerning health, they are 44% and 35% being affected by a physical (i.e., heart disease, lung disease, cancer, high blood pressure, diabetes, severe obesity, and/or autoimmune disease) or a mental (i.e., depressive disorder and/or anxiety disorder) chronic disease, respectively. A monthly consumption of alcohol, cigarettes and/or vape, and cannabis was reported by 72%, 14%, and 18% of participants at baseline, respectively. Lastly, they are 27% and 38% considering their selves and/or someone in their household at a high risk of being infected by- or of complications of COVID-19, respectively.

**Figure 3.**
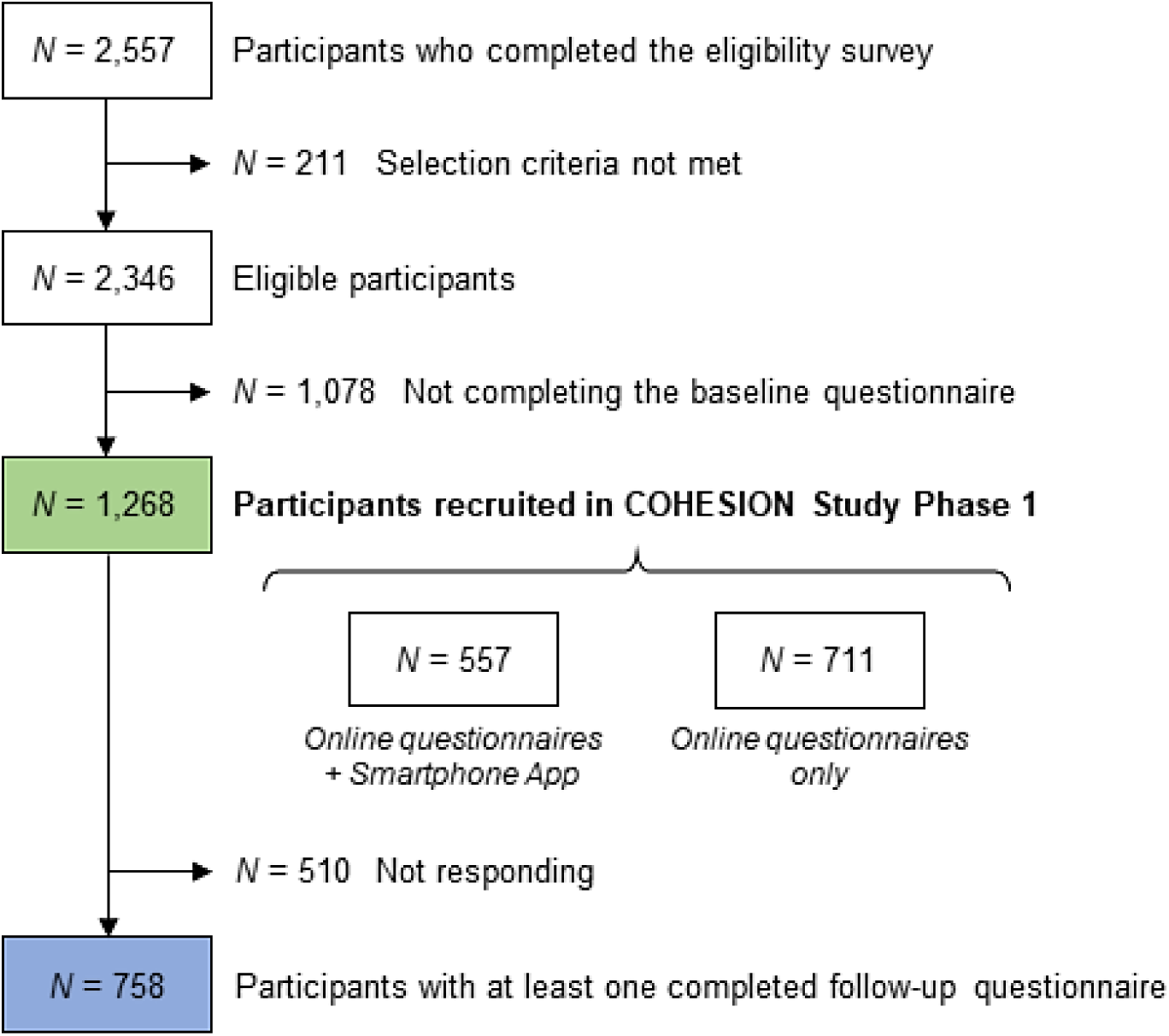
Flow chart of participants’ recruitment to the COHESION Study Phase 1 (June 2020 to July 2021).

**Figure 4.**
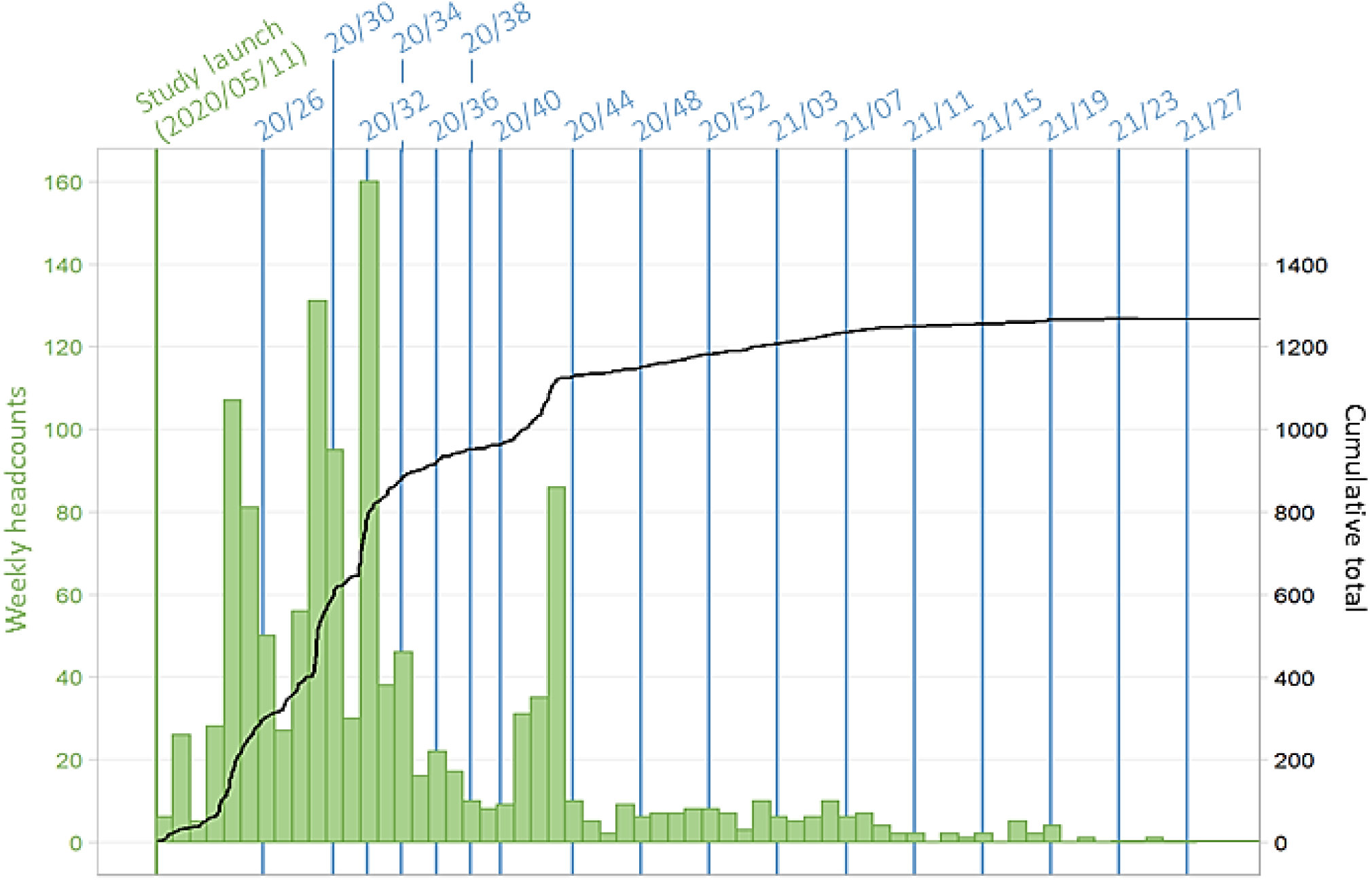
Timeline of recruitment of the COHESION Study Phase 1 participants (*N* = 1,268). Follow-up questionnaire waves are named according to their week and year of release (for instance, “20/26” for the follow-up questionnaire proposed to participants in the 26^th^ week of 2020).

**Figure 5.**
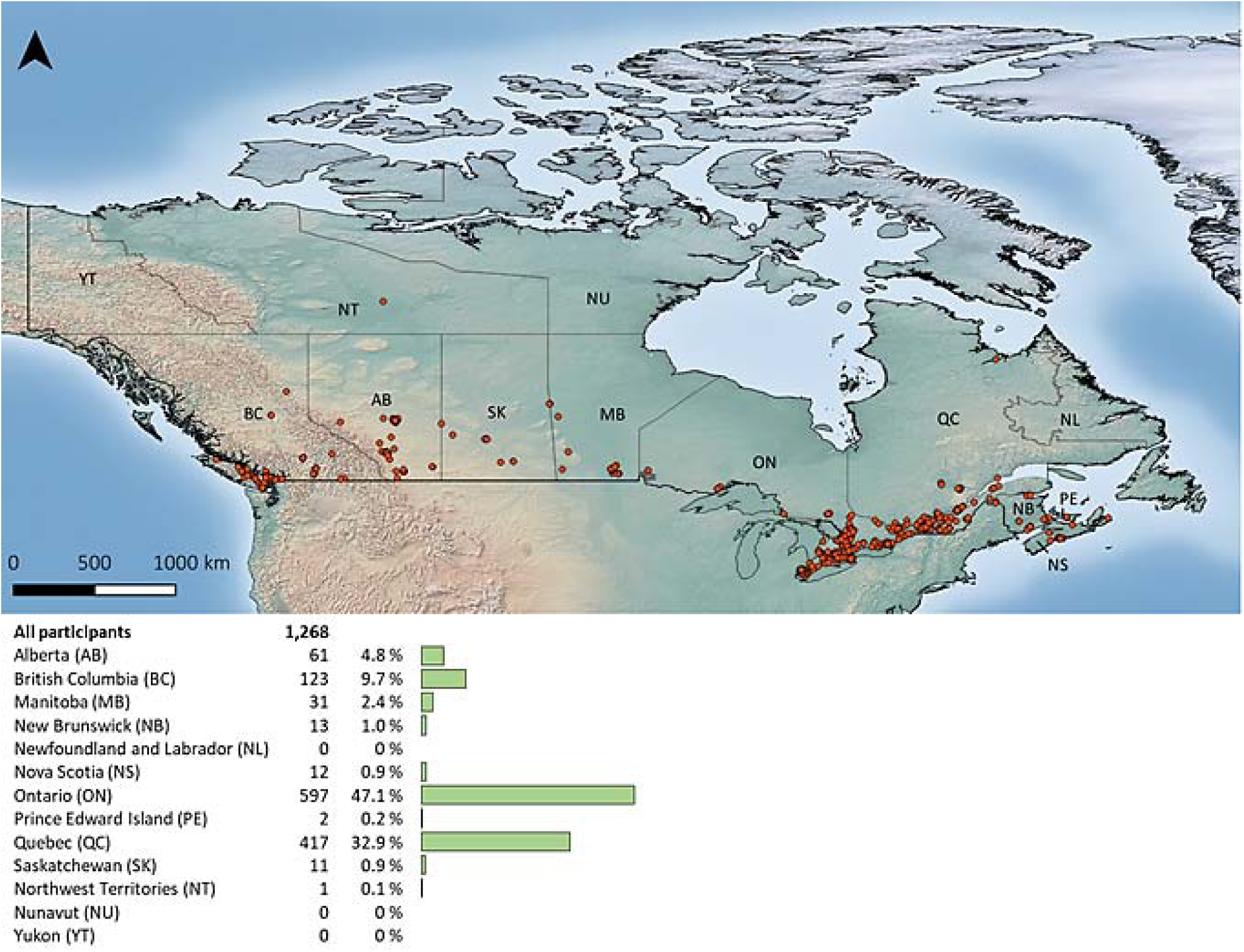
Geographical distribution of COHESION Study Phase 1 participants across Canada (*N* = 1,268).

**Table 2a.** Description of COHESION Study Phase 1 participants, at recruitment (N = 1,268).

| Characteristics | *N* | Percent |
| --- | --- | --- |
| **Demographics** |  |  |
| **Age category** |  |  |
| 15-24 | 103 | 8.1 % |
| 25-34 | 231 | 18.2 % |
| 35-44 | 227 | 17.9 % |
| 45-54 | 230 | 18.1 % |
| 55-64 | 282 | 22.2 % |
| ≥65 | 195 | 15.4 % |
| **Gender** |  |  |
| Woman | 985 | 77.7 % |
| Man | 243 | 19.2 % |
| Other identity | 40 | 3.2 % |
| **Ethnic or cultural group** |  |  |
| White | 1,080 | 85.2 % |
| Other group | 188 | 14.8 % |
| **Born in Canada** |  |  |
| Yes | 1,073 | 84.6 % |
| No | 195 | 15.4 % |
| **Education level** |  |  |
| School degree | 179 | 14.1 % |
| College degree | 278 | 21.9 % |
| University degree | 408 | 32.2 % |
| Post-graduate degree | 393 | 31.0 % |
| Unknown / Not answered | 10 | 0.8 % |
| **Employment** |  |  |
| Employed, full-time | 545 | 43.0 % |
| Employed, part-time | 148 | 11.7 % |
| Self-employed | 36 | 2.8 % |
| Unemployed | 183 | 14.4 % |
| Retired | 245 | 19.3 % |
| On leave / Disabled | 59 | 4.7 % |
| Student | 24 | 1.9 % |
| Other | 28 | 2.2 % |
| **Relationship** |  |  |
| In a relationship / Married | 847 | 66.8 % |
| Single | 282 | 22.2 % |
| Separated / Divorced / Widowed | 135 | 10.6 % |
| Other | 4 | 0.3 % |
| **Household characteristics** |  |  |
| **Household composition** |  |  |
| Alone | 274 | 21.6 % |
| With my family | 482 | 38.0 % |
| With my partner | 451 | 35.6 % |
| Other | 61 | 4.8 % |
| **Children (living at home)** |  |  |
| 0 | 918 | 72.4 % |
| 1 | 155 | 12.2 % |
| 2 | 137 | 10.8 % |
| ≥3 | 58 | 4.6 % |
| **Household annual income** |  |  |
| <\$21,000 | 153 | 12.1 % |
| \$21,000-\$36,000 | 173 | 13.6 % |
| \$36,000-\$48,000 | 141 | 11.1 % |
| \$48,000-\$96,000 | 337 | 26.6 % |
| ≥\$96,000 | 279 | 22.0 % |
| Unknown / Not answered | 185 | 14.6 % |

**Table 2a.** To be continued.
| **Characteristics** | **N** | **Percent** |
| --- | --- | --- |
| **Satisfaction regarding income/needs** |  |  |
| Very well | 453 | 35.7 % |
| Well | 522 | 41.2 % |
| Not well | 271 | 21.4 % |
| Unknown / Not answered | 22 | 1.7 % |
| **Pet ownership** |  |  |
| Yes | 693 | 56.4 % |
| No | 536 | 43.6 % |
| **Housing** |  |  |
| **House tenure** |  |  |
| Owner | 786 | 62.0 % |
| Renter | 382 | 30.1 % |
| Other | 69 | 5.4 % |
| Unknown / Not answered | 31 | 2.4 % |
| **Dwelling type** |  |  |
| House | 833 | 65.7 % |
| Apartment / Condo | 419 | 33.0 % |
| Other | 12 | 0.9 % |
| Unknown / Not answered | 4 | 0.3 % |
| **Private outside space** |  |  |
| Yard or nature | 963 | 78.4 % |
| Balcony or terrace | 225 | 18.3 % |
| No | 41 | 3.3 % |
| **General health** |  |  |
| **Chronic physical disease** |  |  |
| Yes | 557 | 43.9 % |
| No | 688 | 54.3 % |
| Unknown / Not answered | 23 | 1.8 % |
| **Chronic mental disease** |  |  |
| Yes | 441 | 34.8 % |
| No | 794 | 62.6 % |
| Unknown / Not answered | 33 | 2.6 % |
| **Substance use** |  |  |
| **Alcohol <sup>a</sup>** |  |  |
| No | 352 | 27.8 % |
| At least once a month | 245 | 19.3 % |
| At least once a week | 465 | 36.7 % |
| At least once a day | 206 | 16.2 % |
| **Cigarettes and/or vape <sup>a</sup>** |  |  |
| No | 1,095 | 86.4 % |
| At least once a month | 16 | 1.3 % |
| At least once a week | 11 | 0.9 % |
| At least once a day | 146 | 11.5 % |
| **Cannabis <sup>a</sup>** |  |  |
| No | 1,036 | 81.7 % |
| At least once a month | 88 | 6.9 % |
| At least once a week | 57 | 4.5 % |
| At least once a day | 87 | 6.9 % |
| **COVID-19 <sup>b</sup>** |  |  |
| **At a high risk of being infected** |  |  |
| Yes | 327 | 26.6 % |
| No | 902 | 73.4 % |
| **At a high risk of complications** |  |  |
| Yes | 465 | 37.8 % |
| No | 764 | 62.2 % |

<sup>a</sup> During the month before completing baseline questionnaire.<sup>b</sup> Participant and/or someone in his/her household.

**Table 2b.** Residential social and environmental measures (N = 1,268).

| **Indexes** | ***N*** | **Percent** |
| --- | --- | --- |
| **Urbanization degree <sup>a</sup>** |  |  |
| Large center | 891 | 70.4 % |
| Medium center | 102 | 8.1 % |
| Small center | 138 | 10.9 % |
| Rural | 135 | 10.7 % |
| **Surrounding greenness (NDVI) <sup>b</sup>** |  |  |
| <0.28 | 254 | 20.1 % |
| 0.28-0.36 | 259 | 20.5 % |
| 0.36-0.43 | 266 | 21.0 % |
| 0.43-0.51 | 233 | 18.4 % |
| ≥0.51 | 253 | 20.0 % |
| **Material deprivation index <sup>c</sup>** |  |  |
| Quintile 1 | 295 | 24.5 % |
| Quintile 2 | 279 | 23.2 % |
| Quintile 3 | 257 | 21.4 % |
| Quintile 4 | 203 | 16.9 % |
| Quintile 5 | 169 | 14.0 % |
| **Social deprivation index <sup>c</sup>** |  |  |
| Quintile 1 | 162 | 13.5 % |
| Quintile 2 | 162 | 13.5 % |
| Quintile 3 | 199 | 16.5 % |
| Quintile 4 | 251 | 20.9 % |
| Quintile 5 | 429 | 35.7 % |

<sup>a</sup> According to *Statistics Canada* classification, “small”, “medium” and “large” centers correspond to areas including between 1,000 and 29,999, between 30,000 and 99,999, and 100,000 and more inhabitants, respectively, while “rural area” is a residual value gathering all areas located outside population centers (at the four-digit code area level; data 2016) [51].
<sup>b</sup> Quintiles of the growing season Normalized Difference Vegetation Index (NDVI) in the COHESION Study cohort (at the six-digit code area level; data 2019) [52,53].
<sup>c</sup> Quintiles of the distribution over the whole Canadian territory (at the Canadian Census dissemination area level; data 2016) [50].

### 4.2. Prospective follow-up and attrition

Throughout the first year of follow-up, i.e., from May 2020 to July 2021, up to 17 follow-up questionnaires were administered to participants (Fig 4), and 758 (60%) participants completed at least one (Fig 3). Depending on their date of recruitment, participants were contacted between one and seventeen times as part of the prospective follow-up waves (Fig 4), and they filled out in average 29% ± 36% (mean ± SD) of the follow-up questionnaires they received. Among the first year of follow-up, in average 298 ± 68 participants completed the follow-up questionnaire by wave (Fig 6, *see* S1 Table).

**Figure 6.**
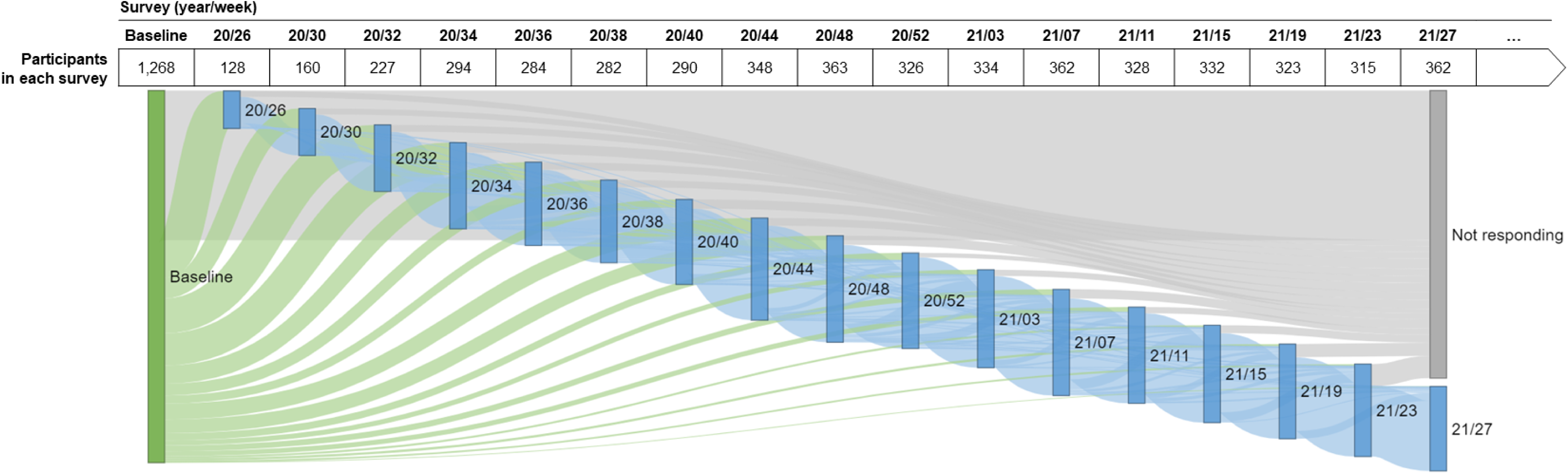
Participation in Phase 1 of the COHESION Study, baseline and follow-ups (June 2020 to July 2021; *N* = 1,268). Follow-up questionnaire waves are named according to their week and year of release (for instance, “20/26” for the follow-up questionnaire proposed to participants in the 26^th^ week of 2020). Green flow shows participants responding for the first time to a follow-up questionnaire, blue flow shows participants responding again to a follow-up questionnaire, and grey flow shows participants not responding to follow-ups.

Attrition rate throughout the Phase 1 follow-up appeared statistically distinct in some specific groups (Table 3). The older the participants were and the higher the education level was, the less the attrition was (*p* <0.001 and *p* = 0.069, respectively); average age was 50 ± 16 and 45 ± 16 (mean ± SD) in people participating or not in the longitudinal follow-up, respectively (*p* <0.001). Attrition appeared higher in employed people and lower in retired ones (*p* <0.001) (Table 3), and also higher in people living in family (*p* = 0.005), with children (*p* = 0.013), with pets (*p* = 0.001), in a house (*p* = 0.001), and not owner (*p* = 0.001) (*see* S2a Table). Lastly, people with a chronic physical disease (*p* = 0.039) or at a high risk of being infected by the COVID-19 (*p* = 0.001) were less prone to participate to the follow-up. No contrast was noticed according to substance use or residential social and environmental components (*see* S2a-b Tables).

**Table 3.** Attrition in the COHESION Study Phase 1, and comparison of participants enrolling or not in the prospective follow-up according to demographics (N = 1,268).

| Characteristics | Participating in<br>follow-ups ($N = 758$) | | Participated in<br>baseline only ($N = 510$) | | *p*-value <sup>a</sup> |
| --- | --- | --- | --- | --- | --- |
|  | *N* | Percent | *N* | Percent |  |
| **Age category** |  |  |  |  | <0.001 |
| 15-24 | 47 | 6.2 % | 56 | 11.0 % |  |
| 25-34 | 120 | 15.8 % | 111 | 21.8 % |  |
| 35-44 | 134 | 17.7 % | 93 | 18.2 % |  |
| 45-54 | 131 | 17.3 % | 99 | 19.4 % |  |
| 55-64 | 183 | 24.1 % | 99 | 19.4 % |  |
| ≥65 | 143 | 18.9 % | 52 | 10.2 % |  |
| **Gender** |  |  |  |  | 0.63 |
| Woman | 591 | 78.0 % | 394 | 77.3 % |  |
| Man | 146 | 19.3 % | 97 | 19.0 % |  |
| Other identity | 21 | 2.8 % | 19 | 3.7 % |  |
| **Ethnic or cultural group** |  |  |  |  | 0.64 |
| White | 649 | 85.6 % | 431 | 84.5 % |  |
| Other group | 109 | 14.4 % | 79 | 15.5 % |  |
| **Born in Canada** |  |  |  |  | 0.12 |
| Yes | 631 | 83.2 % | 442 | 86.7 % |  |
| No | 127 | 16.8 % | 68 | 13.3 % |  |
| **Education level** |  |  |  |  | 0.069 |
| School degree | 94 | 12.4 % | 85 | 16.7 % |  |
| College degree | 169 | 22.3 % | 109 | 21.4 % |  |
| University degree | 235 | 31.0 % | 173 | 33.9 % |  |
| Post-graduate degree | 254 | 33.5 % | 139 | 27.3 % |  |
| Unknown / Not answered | 6 | 0.8 % | 4 | 0.8 % |  |
| **Employment** |  |  |  |  | <0.001 |
| Employed, full-time | 300 | 39.6 % | 245 | 48.0 % |  |
| Employed, part-time | 85 | 11.2 % | 63 | 12.4 % |  |
| Self-employed | 23 | 3.0 % | 13 | 2.5 % |  |
| Unemployed | 99 | 13.1 % | 84 | 16.5 % |  |
| Retired | 192 | 25.3 % | 53 | 10.4 % |  |
| On leave / Disabled | 29 | 3.8 % | 30 | 5.9 % |  |
| Student | 13 | 1.7 % | 11 | 2.2 % |  |
| Other | 17 | 2.2 % | 11 | 2.2 % |  |
| **Relationship** |  |  |  |  | 0.59 |
| In a relationship / Married | 511 | 67.4 % | 336 | 65.9 % |  |
| Single | 160 | 21.1 % | 122 | 23.9 % |  |
| Separated / Divorced / Widowed | 85 | 11.2 % | 50 | 9.8 % |  |
| Other | 2 | 0.3 % | 2 | 0.4 % |  |

<sup>a</sup> Chi-square test (or Fisher exact test for low numbers).

### 4.3. Well-being and mental health

From June 2020 to July 2021, we administered to participants well-being, sleep credit, loneliness, anxiety, depression and psychological distress-related standardized modules 5, 9, 13, 8, 8, and 11 times, respectively (Table 1). During this period, each thematic module was completed at least once by 481 (38%), 630 (50%), 658 (52%), 612 (48%), 612 (48%), and 692 (55%) Phase 1 participants, respectively (*see* S3a-b Tables).

The median of the 5-WHO well-being Index ranged from 44% (interval interquartile, IQR: 24-65; wave 21/27) to 52% (IQR: 32-72; wave 20/52) depending on the wave. Well-being appeared decreasing waves after waves, the median index value from the last two administered waves (21/23 and 21/27, it means 2021, June and July) being significantly lower than the one from the first three ones (20/52, 21/15, and 21/19, it means 2020, December, 2021, April, and 2021, May, respectively) (Fig 7a; *see* S4a Table). The sleep duration appeared stable throughout the first year of the Phase 1 follow-up (mean ± SD: from 7.9 ± 1.3 to 8.0 ± 1.4 hours; waves 21/23 and 20/30, respectively); no change was statistically found. During the prospective follow-up, the median of the UCLA 3-item score of loneliness oscillated between 5 (IQR: 3-6; wave 21/27) and 6 (IQR: 4-7; wave 21/15). These changes were statistically significant, with a feeling of loneliness higher during the period from 2020, October (wave 20/44) to 2021, April (wave 21/15), yet for the wave 20/52, corresponding to the festive season. Moreover, all UCLA 3-item score median values through the follow-up were significantly higher than the UCLA 3-item score median assessed retrospectively for the period prior to the COVID-19 pandemic (4; IQR: 3-6).

**Figure 7a.**
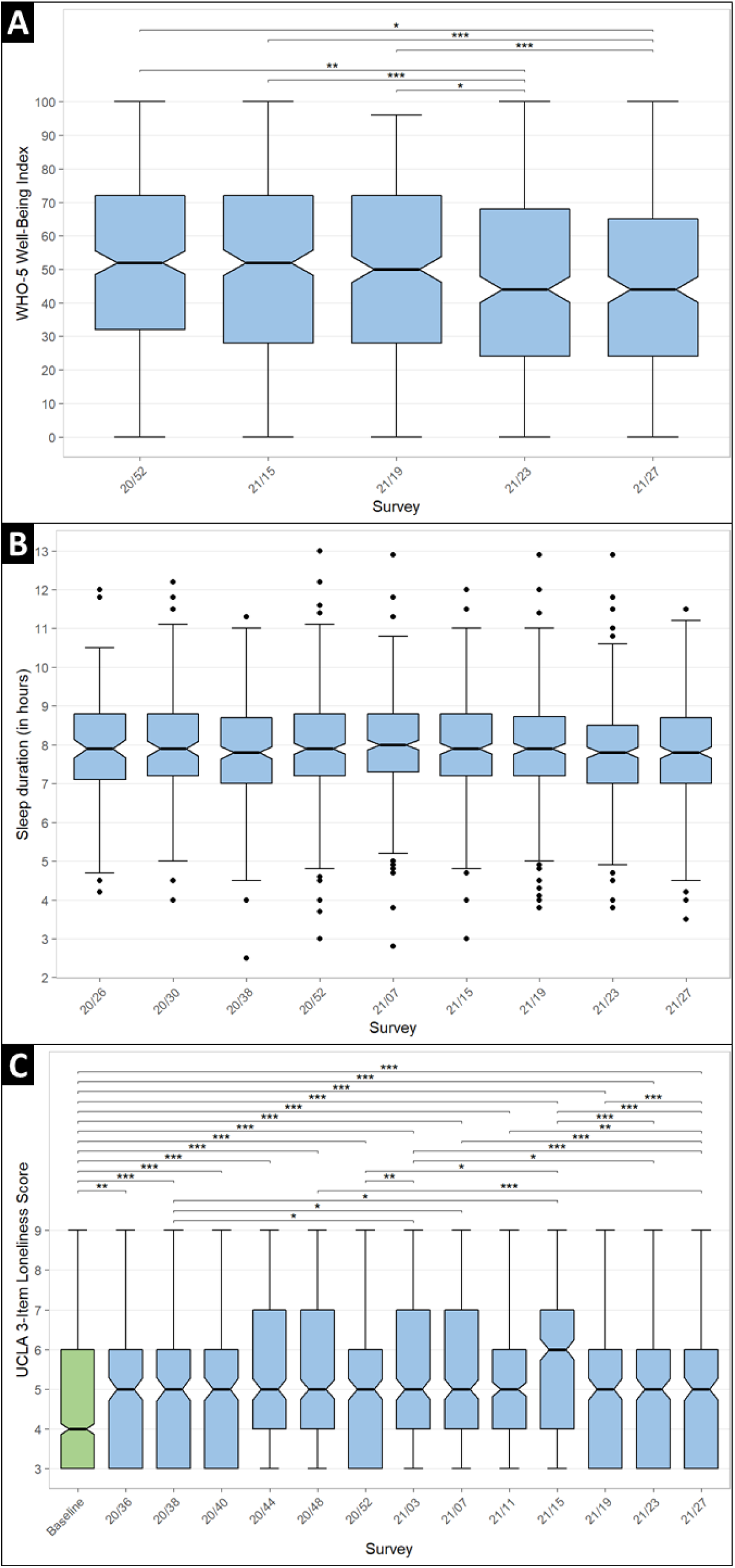
(A) Well-being (WHO-5 Index) level, (B) sleep duration, and (C) feeling of loneliness (UCLA 3-item loneliness score) throughout the first year of prospective follow-up of the COHESION Study Phase 1. Significance of comparison tests, after Bonferroni correction: * <0.05, ** <0.01, *** <0.001. Green: baseline questionnaire; blue: follow-up questionnaire. Follow-up questionnaire waves are named according to their week and year of release (for instance, “20/26” for the follow-up questionnaire proposed to participants in the 26^th^ week of 2020).

Concerning mental health, the first year of follow-up of the Phase 1 showed statistically significant changes between questionnaire waves regarding reporting of anxiety symptoms, depression symptoms, and psychological distress (Fig 7b; *see* S4b Table). Thus, GAD-7 median score ranged between 4 (IQR: 1-7; wave 21/27) and 5 (IQR: 2-10; wave 21/15) and PHQ-9 median score varied from 4 (IQR: 1-7; wave 21/27) and 6 (IQR: 3-9.3; wave 20/30). For these both composite indexes, the median value from the wave 21/27 (2021, July) was the lowest among all waves administered during the first year of follow-up, and some statistically significant contrasts were observed. Lastly, Kessler-6 psychological distress median score oscillated from 4 (IQR: 1-9; wave 20/52) and 5 (IQR: 2-10; wave 20/44); several comparisons between psychological distress median score from the wave 20/52 (2020, December) and other waves were statistically significant.

**Figure 7b.**
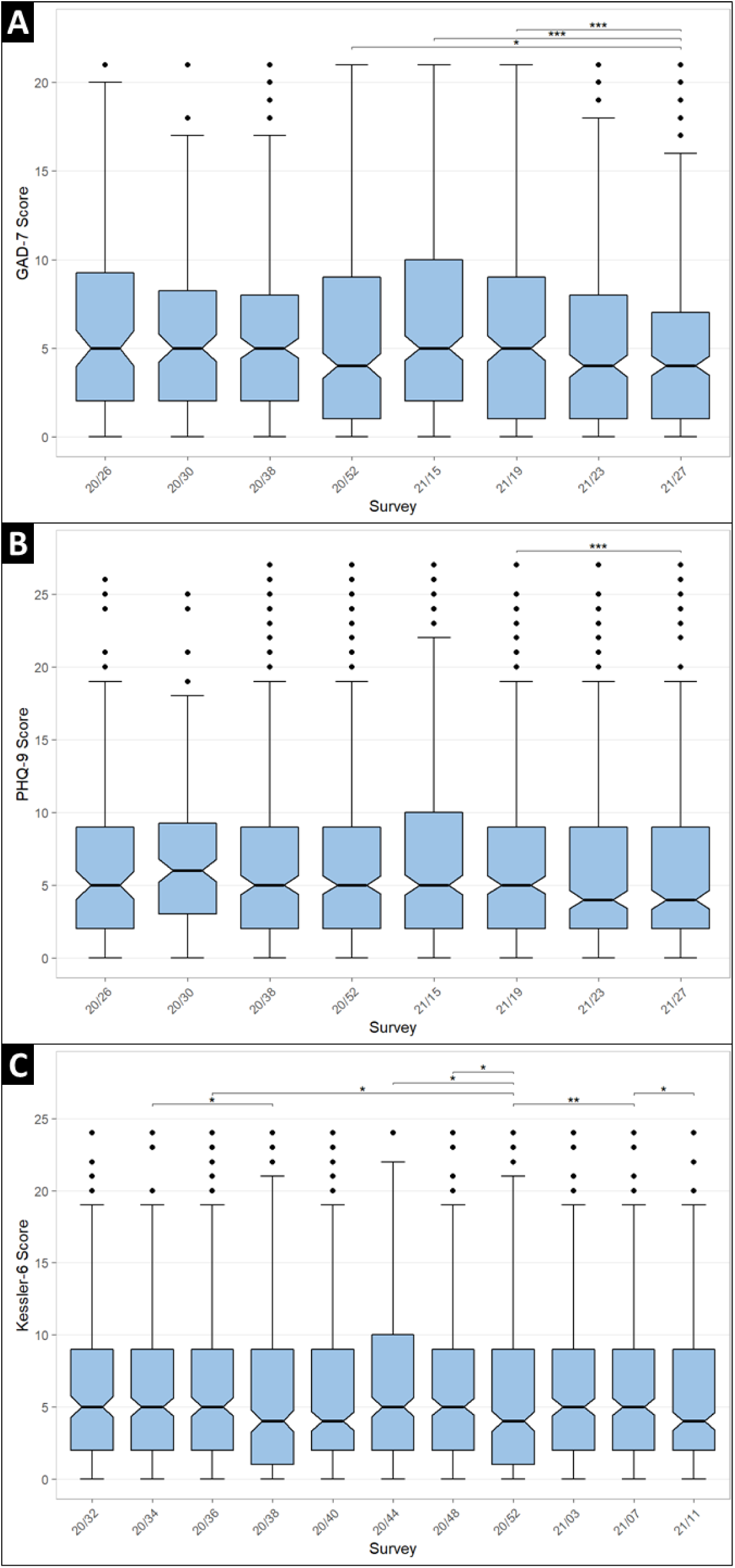
Symptoms of (A) anxiety (7-item General Anxiety Disorder, GAD-7, score), (B) depression (9-item Patient Health Questionnaire, PHQ-9, score), and (C) psychological distress (6-item Kessler Psychological Distress Scale) throughout the first year of prospective follow-up of the COHESION Study Phase 1. Significance of comparison tests, after Bonferroni correction: * <0.05, ** <0.01, *** <0.001. Follow-up questionnaire waves are named according to their week and year of release (for instance, “20/26” for the follow-up questionnaire proposed to participants in the 26^th^ week of 2020

## 5. Discussion

In total, more than 1,200 people enrolled in the Phase 1 of the pan-Canadian population-based COHESION cohort, and about 60% of them participated in the prospective follow-up. The COHESION Study overrepresents women, White and graduated people. The first year of follow-up reveals significant temporal variations in standardized indices of well-being, loneliness, anxiety, depression, and psychological distress. Thus, well-being appeared decreasing waves after waves over the follow-up whereas sleep duration appeared stable throughout. Feeling of loneliness was significantly higher for each follow-up wave compared to the period prior to the COVID-19 pandemic while oscillations in mental health indices were observed over the follow-up timespan. Indices of loneliness and psychological distress were significantly better around the festive period.

### Challenges. lessons learned from Phase 1, and proposed adaptations for Phase 2

Loss to follow-up is an issue in any longitudinal study. In Phase 1, attrition was related to age, education level, employment, housing conditions, and some health conditions, similar to other longitudinal studies [57,58]. To try to reduce attrition in Phase 2 and better retain those that are at greatest risk of loss to follow-up, we have devised the following strategies: 1) Reminders: Our questionnaire platform provides automated reminders for online follow-up survey completion including for EMA; 2) Flexibility: Phase 2 will offer easier participation options, relying on a multi-stage on-boarding process, which includes a short initial 10-min questionnaire and easy-to-sign-up longitudinal follow-ups, with possible ‘light’ or ‘complete’ choices. To increase accessibility, participation and retention (notably with younger participants), we have also optimized the online questionnaire for ease of use through smartphones. 3) Gifts: While enrollment and participation to the COHESION Study are free and voluntary, a lottery of 3 monthly prizes ($100 gift cards) is organized for active participants. In Phase 2, participants who choose the full participation (by completing the optional follow-up questionnaire modules) will be entered twice in the raffle to increase their chance of winning. 4) Improved feedback: In December 2020, an opinion survey was sent to participants to gather feedback on the experience of COHESION Study Phase 1, including on perceived complexity, length, and usability of the questionnaires, interest in surveyed topics, interactions with the mobile application. Suggestions for improvement included better messaging on the purpose of the study, including through regular updates and better recognition of their contribution, and sharing stories to increase the sense of belonging. These elements were considered when re-designing the study for Phase 2, in collaboration with the Center of Excellence on Partnership with Patients and the Public [59].

It is important to include marginalized populations better. We have worked in collaboration with public health partners to identify best recruitment strategies, including by building connections with local partners (e.g., Médecins du Monde, Red Cross, Food banks) that are working directly with various priority groups and have established trust with these individuals and communities [60]. The use of high-precision targeted social media campaigns, provided by partnering consumer research company Potloc Inc., along with continuous monitoring of stratified targets by region, age and gender, should also contribute to optimize sample representation. Now, because the survey uses online technologies that may be a barrier for participation in remote communities and for more marginalized groups, we will also offer computer-assisted phone interviews during working hours through our study helpline.

Finally, as suggested by Fumagalli et al. (2013), promising approaches based on persona principles as those used in marketing strategies might help. Thus, we plan to tailor the content of newsletters, to strengthen the sense of belonging of those most at risk of dropping out, a strategy that has proved effective for re-engaging young and busy people [61]. Despite all these methodological efforts to minimize it, the risk of attrition can hardly be eliminated. One way to account for the potential effect of residual attrition bias is through various analytical strategies, including the use of inverse probability weights for trajectory analyses (e.g., growth curve models) on mental health and well-being.

One critical issue with our cohort that was launched in mid-spring 2020 is the lack of a true pre-pandemic baseline. This can partly be circumvented with retrospective questions, but as time goes by, the recall bias increases, particularly for subjective mental health measures [62]. After first using specific ‘pre-pandemic’ reference questions, a strategy we adopted, from fall 2020 onwards, to minimize the cognitive effort and bias, was to use a combination of broader questions about psychological changes since the pandemic outbreak (improvement, deterioration, no change) and very factual questions for which recall bias is expected lower (i.e., use of mental health services and substance use in the pre-pandemic year). Despite these adaptations, the risk of recall bias remains, meaning that interpretation about change from pre-pandemic measures should be done with caution.

An advantage of a prospective cohort design is the potential for adapting surveys to new or unforeseen circumstances, including evolving priorities of public health authorities. As months passed by and the situation evolved, we developed and administered new modules to address such needs, including on substance use, a priority identified by our partner the Public Health Agency of Canada, or on parent-related stress during the back-to-school periods. Repeated surveys also provide the opportunity for timely dissemination of findings. To facilitate dissemination and use of data by our public health partners, we developed an online dashboard for real-time monitoring of key indicators (www.cohesionstudy.ca/dashboard). Throughout phase 2 of COHESION, we will continue adapting our survey content with timely themes, helping uncovering key pathways linking individual trajectories, environmental contexts, and health and equity outcomes.

An important contribution of this project is to offer both a pan-Canadian portrait while allowing local oversampling and providing very contextualized information. An example of local adaptation is planned for public health territory of CIUSSS Nord-de-l’Île-de-Montréal (NIM) in Québec [63], with a target local sample of 1,000 participants. Data will be used to support the co-construction of tailored local intersectoral interventions towards mental health and more broadly to support activities towards health equity as part of a longer-term pandemic recovery strategy.

While COHESION Study Phase 1 aimed to understand the mechanisms linking residential living conditions (built environment, surrounding greenness, neighborhood deprivation) to differential trajectories of mental health and well-being since the outbreak of the pandemic, Phase 2 may also provide decision support for public health authorities across Canada. This contextualized information on a potential representative sample of general population would be particularly important for policy makers in the pandemic recovery period where it will be crucial to address health inequities related to income, housing, daily mobility and social interactions, intimate partner violence, childhood, access to food and health care, and racism exacerbated over the past two years.

## 6. Conclusion

Real-time monitoring and evaluation of the unintended consequences on mental health and health inequities of the pandemic is essential for shaping and adapting effective public health policies and programs targeting contextual living conditions (e.g., pedestrianization of streets, securing access to parks, housing renovation programs, permanent supportive housing programs, neighborhood greening program). We will benefit from the support of the Uni-Cité Collaboratory [64] which specializes in science to policy approaches – to equip research teams and cities with tools to better incorporate science into urban public policy. Our study will provide a comprehensive portrait of the key pathways of the COVID-19 impacts on mental health and well-being across Canada. Our flexible infrastructure will ensure that we can adjust to local needs and to the evolving situation from pandemic to post-pandemic recovery.

## Data Availability

All relevant data are within the manuscript and its Supporting Information files. A public dashboard is also available at: https://cohesionstudy.ca/dashboard/

## 7. Acknowledgments

The authors would like to warmly thank Dre. Kate Zinszer (*Centre de Recherche en Santé Publique*, Montreal, QC, Canada) for her crucial involvement in the foundation of this project; Alexandra Otis (*CIUSSS Nord-de-l’Ile-de-Montréal*, Montreal, QC, Canada) and Marina Najjar (*Centre de Recherche en Santé Publique*, Montreal, QC, Canada) for their decisive coordination work in the Phase 1 COHESION study; Alexie Kim, Salma Sahil, and Andréanne Fortin for their assistance in study’s communication; Alexandre Naud for his help with the VERITAS questionnaire; and Catherine Grenon for her help in analyzing the verbatims of the completed surveys. NDVI and MSDI metrics, indexed to DMTI Spatial Inc. postal codes, were provided by the *Canadian Urban Environmental Health Research Consortium* (CANUE).

