## Supplemental material for "How is the COVID-19 pandemic impacting our life, mental health, and well-being? Design and preliminary findings of the pan-Canadian longitudinal COHESION Study"

**Supporting information**

4 tables and 2 figures (in color).

**Table S1.** Participation in the COHESION Study Phase 1 throughout the first year of prospective follow-up (June 2020 to July 2021): detailed statistics (*N* = 1,268).

**Next completed survey (year/week)**

|  |  | **Survey (year/week)** | | |  |  |  |  |  |  |  |  |  |  |  |  |  |  |  |
| --- | --- | --- | --- | --- | --- | --- | --- | --- | --- | --- | --- | --- | --- | --- | --- | --- | --- | --- | --- |
|  |  | **Baseline** | **20/26** | **20/30** | **20/32** | **20/34** | **20/36** | **20/38** | **20/40** | **20/44** | **20/48** | **20/52** | **21/03** | **21/07** | **21/11** | **21/15** | **21/19** | **21/23** | **21/27** |
| **Participants** |  | **1,268** | **128** | **160** | **227** | **294** | **284** | **282** | **290** | **348** | **363** | **326** | **334** | **362** | **328** | **332** | **323** | **315** | **362** |
| **20/26** |  | 128  (10.1 %) |  |  |  |  |  |  |  |  |  |  |  |  |  |  |  |  |  |
| **20/30** |  | 70  (5.5 %) | 90  (70.3 %) |  |  |  |  |  |  |  |  |  |  |  |  |  |  |  |  |
| **20/32** |  | 119  (9.4 %) | 9  (7.0 %) | 99  (61.9 %) |  |  |  |  |  |  |  |  |  |  |  |  |  |  |  |
| **20/34** |  | 110  (8.7 %) | 3  (2.3 %) | 29  (18.1 %) | 152  (67.0 %) |  |  |  |  |  |  |  |  |  |  |  |  |  |  |
| **20/36** |  | 62  (4.9 %) | 1  (0.8 %) | 3  (1.9 %) | 35  (15.4 %) | 183  (62.2 %) |  |  |  |  |  |  |  |  |  |  |  |  |  |
| **20/38** |  | 40  (3.2 %) | - | 3  (1.9 %) | 8  (3.5 %) | 51  (17.3 %) | 180  (63.4 %) |  |  |  |  |  |  |  |  |  |  |  |  |
| **20/40** |  | 28  (2.2 %) | - | - | 3  (1.3 %) | 14  (4.8 %) | 53  (18.7 %) | 192  (68.1 %) |  |  |  |  |  |  |  |  |  |  |  |
| **20/44** |  | 504  (4.3 %) | - | 1  (0.6 %) | - | 5  (1.7 %) | 19  (6.7 %) | 45  (16.0 %) | 224  (77.2 %) |  |  |  |  |  |  |  |  |  |  |
| **20/48** |  | 47  (3.7 %) | 2  (1.6 %) | - | 2  (0.9 %) | 2  (0.7 %) | 2  (0.7 %) | 8  (2.8 %) | 19  (6.6 %) | 281  (80.7 %) |  |  |  |  |  |  |  |  |  |
| **20/52** |  | 24  (1.9 %) | - | 2  (1.2 %) | 1  (0.4 %) | 1  (0.3 %) | 1  (0.4 %) | 6  (2.1 %) | 7  (2.4 %) | 15  (4.3 %) | 269  (74.1 %) |  |  |  |  |  |  |  |  |
| **21/03** |  | 17  (1.3 %) | 1  (0.8 %) | - | 1  (0.4 %) | - | 2  (0.7 %) | 3  (1.1 %) | 3  (1.0 %) | 9  (2.6 %) | 35  (9.6 %) | 263  (80.7 %) |  |  |  |  |  |  |  |
| **21/07** |  | 20  (1.6 %) | - | 1  (0.6 %) | - | 2  (0.7 %) | - | 1  (0.4 %) | 2  (0.7 %) | 6  (1.7 %) | 12  (3.3 %) | 31  (9.5 %) | 287  (85.9 %) |  |  |  |  |  |  |
| **21/11** |  | 17  (1.3 %) | - | - | 1  (0.4 %) | - | - | 2  (0.7 %) | - | 3  (0.9 %) | 3  (0.8 %) | 8  (2.5 %) | 18  (5.4 %) | 276  (76.2 %) |  |  |  |  |  |
| **21/15** |  | 7  (0.6 %) | - | 1  (0.6 %) | - | 1  (0.3 %) | 1  (0.4 %) | - | 1  (0.3 %) | 6  (1.7 %) | 2  (0.6 %) | 4  (1.2 %) | 6  (1.8 %) | 35  (9.7 %) | 268  (81.7 %) |  |  |  |  |
| **21/19** |  | 8  (0.6 %) | 1  (0.8 %) | 1  (0.6 %) | - | 1  (0.3 %) | - | - | 1  (0.3 %) | 1  (0.3 %) | 6  (1.7 %) | 3  (0.9 %) | - | 9  (2.5 %) | 22  (6.7 %) | 270  (81.3 %) |  |  |  |
| **21/23** |  | 6  (0.5 %) | - | - | 1  (0.4 %) | - | 1  (0.4 %) | 1  (0.4 %) | 1  (0.3 %) | - | 1  (0.3 %) | 1  (0.3 %) | 1  (0.3 %) | 8  (2.2 %) | 11  (3.4 %) | 32  (9.6 %) | 251  (77.7 %) |  |  |
| **21/27** |  | 1  (0.1 %) | 1  (0.8 %) | - | - | - | - | - | 1  (0.3 %) | 1  (0.3 %) | 4  (1.1 %) | 1  (0.3 %) | 1  (0.3 %) | 3  (0.8 %) | 6  (1.8 %) | 3  (0.9 %) | 27  (8.4 %) | 239  (75.9 %) |  |
| **Not**  **responding** |  | 510  (40.2 %) | 20  (15.6 %) | 20  (12.5 %) | 23  (10.1 %) | 34  (11.6 %) | 25  (8.8 %) | 24  (8.5 %) | 31  (10.7 %) | 26  (7.5 %) | 31  (8.5 %) | 15  (4.6 %) | 21  (6.3 %) | 31  (8.6 %) | 21  (6.4 %) | 27  (8.1 %) | 45  (13.9 %) | 76  (24.1 %) |  |

Headcounts correspond to flows of participants having completed the questionnaire in header of column then the questionnaire in header of row; percentages correspond to headcounts by the total number of participants having completed the questionnaire in header of column. Follow-up questionnaire waves are named according to their week and year of release (for instance, “20/26” for the follow-up questionnaire proposed to participants in the 26^th^ week of 2020).

**Table S2a.** Comparison of participants enrolling or not in the prospective follow-up according to household characteristics, housing, health, substance use, and COVID-19-related risks (*N* = 1,268).

|  | **Participating in  follow-ups (*N* = 758)** | | **Participated in baseline only (*N* = 510)** | |  |
| --- | --- | --- | --- | --- | --- |
| **Characteristics** | ***N*** | **Percent** | ***N*** | **Percent** | ***p*-value ^a^** |
| **Household characteristics** |  |  |  |  |  |
| **Household composition** |  |  |  |  | 0.005 |
| Alone | 175 | 23.1 % | 99 | 19.4 % |  |
| With my family | 258 | 34.0 % | 224 | 43.9 % |  |
| With my partner | 287 | 37.9 % | 164 | 32.2 % |  |
| Other | 38 | 5.0 % | 23 | 4.5 % |  |
| **Children (living at home)** |  |  |  |  | 0.013 |
| 0 | 559 | 73.7 % | 359 | 70.4 % |  |
| 1 | 81 | 10.7 % | 74 | 14.5 % |  |
| 2 | 91 | 12.0 % | 46 | 9.0 % |  |
| ≥3 | 27 | 3.6 % | 31 | 6.1 % |  |
| **Household annual income** |  |  |  |  | 0.96 |
| <$21,000 | 90 | 11.9 % | 63 | 12.4 % |  |
| $21,000-$36,000 | 106 | 14.0 % | 67 | 13.1 % |  |
| $36,000-$48,000 | 86 | 11.3 % | 55 | 10.8 % |  |
| $48,000-$96,000 | 206 | 27.2 % | 131 | 25.7 % |  |
| ≥$96,000 | 162 | 21.4 % | 117 | 22.9 % |  |
| Unknown / Not answered | 108 | 14.2 % | 77 | 15.1 % |  |
| **Satisfaction regarding income/needs** |  |  |  |  | 0.11 |
| Very well | 288 | 38.0 % | 165 | 32.4 % |  |
| Well | 307 | 40.5 % | 215 | 42.2 % |  |
| Not well | 153 | 20.2 % | 118 | 23.1 % |  |
| Unknown / Not answered | 10 | 1.3 % | 12 | 2.4 % |  |
| **Pet ownership** |  |  |  |  | 0.001 |
| Yes | 389 | 52.4 % | 304 | 62.4 % |  |
| No | 353 | 47.6 % | 183 | 37.6 % |  |
| **Housing** |  |  |  |  |  |
| **House tenure** |  |  |  |  | 0.001 |
| Owner | 486 | 64.1 % | 300 | 58.8 % |  |
| Renter | 231 | 30.5 % | 151 | 29.6 % |  |
| Other | 30 | 4.0 % | 39 | 7.6 % |  |
| Unknown / Not answered | 11 | 1.5 % | 20 | 3.9 % |  |
| **Dwelling type** |  |  |  |  | 0.017 |
| House | 488 | 64.4 % | 345 | 67.6 % |  |
| Apartment / Condo | 265 | 35.0 % | 154 | 30.2 % |  |
| Other | 5 | 0.7 % | 7 | 1.4 % |  |
| Unknown / Not answered | 0 | 0 % | 4 | 0.8 % |  |
| **Private outside space** |  |  |  |  | 0.19 |
| Yard or nature | 570 | 76.8 % | 393 | 80.7 % |  |
| Balcony or terrace | 143 | 19.3 % | 82 | 16.8 % |  |
| No | 29 | 3.9 % | 12 | 2.5 % |  |
| **General health** |  |  |  |  |  |
| **Chronic physical disease** |  |  |  |  | 0.039 |
| Yes | 330 | 43.5 % | 227 | 44.5 % |  |
| No | 420 | 55.4 % | 268 | 52.5 % |  |
| Unknown / Not answered | 8 | 1.1 % | 15 | 2.9 % |  |
| **Chronic mental disease** |  |  |  |  | 0.23 |
| Yes | 251 | 33.1 % | 190 | 37.3 % |  |
| No | 489 | 64.5 % | 305 | 59.8 % |  |
| Unknown / Not answered | 18 | 2.4 % | 15 | 2.9 % |  |

**Table S2a.** To be continued.

|  | **Participating in  follow-ups (*N* = 758)** | | **Participated in baseline only (*N* = 510)** | |  |
| --- | --- | --- | --- | --- | --- |
| **Characteristics** | ***N*** | **Percent** | ***N*** | **Percent** | ***p*-value ^a^** |
| **Substance use** |  |  |  |  |  |
| **Alcohol ^b^** |  |  |  |  | 0.16 |
| No | 199 | 26.3 % | 153 | 30.0 % |  |
| At least once a month | 148 | 19.5 % | 97 | 19.0 % |  |
| At least once a week | 295 | 38.9 % | 170 | 33.3 % |  |
| At least once a day | 116 | 15.3 % | 90 | 17.6 % |  |
| **Cigarettes and/or vape ^b^** |  |  |  |  | 0.51 |
| No | 661 | 87.2 % | 434 | 85.1 % |  |
| At least once a month | 11 | 1.5 % | 5 | 1.0 % |  |
| At least once a week | 6 | 0.8 % | 5 | 1.0 % |  |
| At least once a day | 80 | 10.6 % | 66 | 12.9 % |  |
| **Cannabis ^b^** |  |  |  |  | 0.20 |
| No | 613 | 80.9 % | 423 | 82.9 % |  |
| At least once a month | 62 | 8.2 % | 26 | 5.1 % |  |
| At least once a week | 33 | 4.4 % | 24 | 4.7 % |  |
| At least once a day | 50 | 6.6 % | 37 | 7.3 % |  |
| **COVID-19 ^c^** |  |  |  |  |  |
| **At a high risk of being infected** |  |  |  |  | 0.001 |
| Yes | 172 | 23.2 % | 155 | 31.8 % |  |
| No | 570 | 76.8 % | 332 | 68.2 % |  |
| **At a high risk of complications** |  |  |  |  | 0.35 |
| Yes | 289 | 38.9 % | 176 | 36.1 % |  |
| No | 453 | 61.1 % | 311 | 63.9 % |  |

^a^ Chi-square test (or Fisher exact test for low numbers).

^b^ During the month before completing baseline questionnaire.

^c^ Participant and/or someone in his/her household.

**Table S2b.** Comparison of participants enrolling or not in the prospective follow-up according to residential social and environmental measures (*N* = 1,268).

|  | **Participating in  follow-ups (*N* = 758)** | | **Participated in baseline only (*N* = 510)** | |  |
| --- | --- | --- | --- | --- | --- |
| **Characteristics** | ***N*** | **Percent** | ***N*** | **Percent** | ***p*-value ^a^** |
| **Urbanization degree ^b^** |  |  |  |  | 0.138 |
| Large center | 547 | 72.2 % | 344 | 67.7 % |  |
| Medium center | 55 | 7.3 % | 47 | 9.3 % |  |
| Small center | 85 | 11.2 % | 53 | 10.4 % |  |
| Rural | 71 | 9.4 % | 64 | 12.6 % |  |
| **Surrounding greenness (NDVI) ^c^** |  |  |  |  | 0.549 |
| <0.28 | 154 | 20.3 % | 100 | 19.7 % |  |
| 0.28-0.36 | 160 | 21.1 % | 99 | 19.5 % |  |
| 0.36-0.43 | 149 | 19.7 % | 117 | 23.0 % |  |
| 0.43-0.51 | 146 | 19.3 % | 87 | 17.1 % |  |
| ≥0.51 | 148 | 19.6 % | 105 | 20.7 % |  |
| **Material deprivation index ^d^** |  |  |  |  | 0.532 |
| Quintile 1 | 178 | 24.6 % | 117 | 24.4 % |  |
| Quintile 2 | 177 | 24.5 % | 102 | 21.2 % |  |
| Quintile 3 | 147 | 20.3 % | 110 | 22.9 % |  |
| Quintile 4 | 125 | 17.3 % | 78 | 16.2 % |  |
| Quintile 5 | 96 | 13.3 % | 73 | 15.2 % |  |
| **Social deprivation index ^d^** |  |  |  |  | 0.492 |
| Quintile 1 | 88 | 12.2 % | 74 | 15.4 % |  |
| Quintile 2 | 99 | 13.7 % | 63 | 13.1 % |  |
| Quintile 3 | 116 | 16.0 % | 83 | 17.3 % |  |
| Quintile 4 | 156 | 21.6 % | 95 | 19.8 % |  |
| Quintile 5 | 264 | 36.5 % | 165 | 34.4 % |  |

^a^ Chi-square test (or Fisher exact test for low numbers).

^b^ According to *Statistics Canada* classification, “small”, “medium” and “large” centers correspond to areas including between 1,000 and 29,999, between 30,000 and 99,999, and 100,000 and more inhabitants, respectively, while “rural area” is a residual value gathering all areas located outside population centers (at the four-digit code area level; data 2016) [51].

^c^ Quintiles of the growing season Normalized Difference Vegetation Index (NDVI) in the COHESION Study cohort (at the six-digit code area level; data 2019) [52,53].

^d^ Quintiles of the distribution over the whole Canadian territory (at the Canadian Census dissemination area level; data 2016) [50].

**Table S3a.** Participation to the thematic questionnaire modules on well-being, sleep credit, and loneliness in the COHESION Study Phase 1 throughout the first year of prospective follow-up (June 2020 to July 2021, *N* = 1,268).

|  | | **Participants** | |  |  | |
| --- | --- | --- | --- | --- | --- | --- |
| **Count of completed  follow-up surveys** | | ***N*** | **Percent** | **Cumulative N** ^a^ | | **Cumulative percent** ^a^ |
| **Well-being** |  | | | | | |
| 0 | | 787 | 62.1 % | – | | – |
| 1 | | 114 | 9.0 % | 481 | | 38.0 % |
| 2 | | 67 | 5.3 % | 367 | | 29.0 % |
| 3 | | 44 | 3.5 % | 300 | | 23.7 % |
| 4 | | 77 | 6.1 % | 256 | | 20.2 % |
| 5 | | 179 | 14.1 % | 179 | | 14.1 % |
| **Sleep credit** |  | | | | | |
| 0 | | 638 | 50.3 % | – | | – |
| 1 | | 153 | 12.1 % | 630 | | 49.7 % |
| 2 | | 100 | 7.9 % | 477 | | 37.6 % |
| 3 | | 56 | 4.4 % | 377 | | 29.7 % |
| 4 | | 46 | 3.6 % | 321 | | 25.3 % |
| 5 | | 50 | 3.9 % | 275 | | 21.7 % |
| 6 | | 87 | 6.9 % | 225 | | 17.8 % |
| 7 | | 86 | 6.8 % | 138 | | 10.9 % |
| 8 | | 31 | 2.4 % | 52 | | 4.1 % |
| 9 | | 21 | 1.7 % | 21 | | 1.7 % |
| **Loneliness** |  | | | | | |
| 0 | | 610 | 48.1 % | – | | – |
| 1 | | 114 | 9.0 % | 658 | | 52.1 % |
| 2 | | 81 | 6.4 % | 544 | | 43.1 % |
| 3 | | 50 | 3.9 % | 463 | | 36.7 % |
| 4 | | 46 | 3.6 % | 413 | | 32.8 % |
| 5 | | 34 | 2.7 % | 367 | | 29.2 % |
| 6 | | 38 | 3.0 % | 333 | | 26.5 % |
| 7 | | 34 | 2.7 % | 295 | | 23.5 % |
| 8 | | 26 | 2.1 % | 261 | | 20.8 % |
| 9 | | 26 | 2.1 % | 235 | | 18.7 % |
| 10 | | 40 | 3.2 % | 209 | | 16.6 % |
| 11 | | 40 | 3.2 % | 169 | | 13.4 % |
| 12 | | 51 | 4.0 % | 129 | | 10.2 % |
| 13 | | 78 | 6.2 % | 78 | | 6.2 % |

^a^ I.e., headcount and percentage of participants having completed at least the corresponding number of follow-ups.

**Table S3b.** Participation to the thematic questionnaire modules on anxiety symptoms, depression symptoms, and psychological distress in the COHESION Study Phase 1 throughout the first year of prospective follow-up (June 2020 to July 2021, *N* = 1,268).

|  | **Participants** | |  |  | |
| --- | --- | --- | --- | --- | --- |
| **Count of completed  follow-up surveys** | ***N*** | **Percent** | **Cumulative N** ^a^ | | **Cumulative percent** ^a^ |
| **Anxiety symptoms** | | | | | |
| 0 | 656 | 51.7 % | – | | – |
| 1 | 163 | 12.9 % | 612 | | 48.4 % |
| 2 | 101 | 8.0 % | 449 | | 35.5 % |
| 3 | 53 | 4.2 % | 348 | | 27.5 % |
| 4 | 68 | 5.4 % | 295 | | 23.3 % |
| 5 | 79 | 6.2 % | 227 | | 17.9 % |
| 6 | 96 | 7.6 % | 148 | | 11.7 % |
| 7 | 30 | 2.4 % | 52 | | 4.1 % |
| 8 | 22 | 1.7 % | 22 | | 1.7 % |
| **Depression symptoms** | | | | | |
| 0 | 656 | 51.7 % | – | | – |
| 1 | 163 | 12.9 % | 612 | | 48.4 % |
| 2 | 101 | 8.0 % | 449 | | 35.5 % |
| 3 | 53 | 4.2 % | 348 | | 27.5 % |
| 4 | 68 | 5.4 % | 295 | | 23.3 % |
| 5 | 79 | 6.2 % | 227 | | 17.9 % |
| 6 | 96 | 7.6 % | 148 | | 11.7 % |
| 7 | 30 | 2.4 % | 52 | | 4.1 % |
| 8 | 22 | 1.7 % | 22 | | 1.7 % |
| **Psychological distress** | | | | | |
| 0 | 576 | 45.4 % | – | | – |
| 1 | 144 | 11.4 % | 692 | | 54.7 % |
| 2 | 102 | 8.0 % | 548 | | 43.3 % |
| 3 | 59 | 4.7 % | 446 | | 35.3 % |
| 4 | 58 | 4.6 % | 387 | | 30.6 % |
| 5 | 48 | 3.8 % | 329 | | 26.0 % |
| 6 | 46 | 3.6 % | 281 | | 22.2 % |
| 7 | 40 | 3.2 % | 235 | | 18.6 % |
| 8 | 34 | 2.7 % | 195 | | 15.4 % |
| 9 | 49 | 3.9 % | 161 | | 12.7 % |
| 10 | 60 | 4.7 % | 112 | | 8.8 % |
| 11 | 52 | 4.1 % | 52 | | 4.1 % |

^a^ I.e., headcount and percentage of participants having completed at least the corresponding number of follow-ups.

**Table S4a.** WHO-5 Index, sleep duration, and UCLA 3-item loneliness score throughout the first year of prospective follow-up of the COHESION Study Phase 1: detailed statistics.

| **Survey** | ***N*** | **Min** | **Q1** | **Q2** | **Q3** | **Max** | **Mean ± SD** |
| --- | --- | --- | --- | --- | --- | --- | --- |
| **Well-being (WHO-5 Index; in %)** | | | | | | | |
| 20/52 | 326 | 0 | 32.0 | 52.0 | 72.0 | 100 | 51.7 ± 25.0 |
| 21/15 | 332 | 0 | 28.0 | 52.0 | 72.0 | 100 | 52.4 ± 25.1 |
| 21/19 | 322 | 0 | 28.0 | 50.0 | 72.0 | 96 | 50.5 ± 24.9 |
| 21/23 | 315 | 0 | 24.0 | 44.0 | 68.0 | 100 | 47.4 ± 25.8 |
| 21/27 | 288 | 0 | 24.0 | 44.0 | 65.0 | 100 | 46.5 ± 24.9 |
| **Sleep duration (in hours)** | | | | | | | |
| 20/26 | 127 | 4.2 | 7.1 | 7.9 | 8.8 | 12.0 | 7.9 ± 1.3 |
| 20/30 | 160 | 4.0 | 7.2 | 7.9 | 8.8 | 12.2 | 8.0 ± 1.4 |
| 20/38 | 280 | 2.5 | 7.0 | 7.8 | 8.7 | 11.3 | 7.8 ± 1.3 |
| 20/52 | 322 | 3.0 | 7.2 | 7.9 | 8.8 | 13.0 | 7.9 ± 1.4 |
| 21/07 | 361 | 2.8 | 7.3 | 8.0 | 8.8 | 12.9 | 8.0 ± 1.3 |
| 21/15 | 329 | 3.0 | 7.2 | 7.9 | 8.8 | 12.0 | 7.9 ± 1.3 |
| 21/19 | 316 | 3.8 | 7.2 | 7.9 | 8.7 | 12.9 | 7.9 ± 1.3 |
| 21/23 | 311 | 3.8 | 7.0 | 7.8 | 8.5 | 12.9 | 7.9 ± 1.3 |
| 21/27 | 283 | 3.5 | 7.0 | 7.8 | 8.7 | 11.5 | 7.8 ± 1.3 |
| **Loneliness (UCLA 3-item score)** | | | | | | | |
| Baseline | 1,268 | 3 | 3.0 | 4.0 | 6.0 | 9 | 4.7 ± 1.8 |
| 20/36 | 281 | 3 | 3.0 | 5.0 | 6.0 | 9 | 5.1 ± 1.9 |
| 20/38 | 275 | 3 | 3.0 | 5.0 | 6.0 | 9 | 5.2 ± 1.9 |
| 20/40 | 287 | 3 | 3.0 | 5.0 | 6.0 | 9 | 5.1 ± 2.0 |
| 20/44 | 341 | 3 | 4.0 | 5.0 | 7.0 | 9 | 5.4 ± 2.0 |
| 20/48 | 363 | 3 | 4.0 | 5.0 | 7.0 | 9 | 5.5 ± 1.9 |
| 20/52 | 326 | 3 | 3.0 | 5.0 | 6.0 | 9 | 5.2 ± 2.0 |
| 21/03 | 334 | 3 | 4.0 | 5.0 | 7.0 | 9 | 5.4 ± 1.9 |
| 21/07 | 362 | 3 | 4.0 | 5.0 | 7.0 | 9 | 5.5 ± 2.0 |
| 21/11 | 328 | 3 | 4.0 | 5.0 | 6.0 | 9 | 5.4 ± 1.9 |
| 21/15 | 332 | 3 | 4.0 | 6.0 | 7.0 | 9 | 5.6 ± 1.9 |
| 21/19 | 322 | 3 | 3.0 | 5.0 | 6.0 | 9 | 5.3 ± 1.9 |
| 21/23 | 315 | 3 | 3.0 | 5.0 | 6.0 | 9 | 5.2 ± 2.0 |
| 21/27 | 288 | 3 | 3.0 | 5.0 | 6.0 | 9 | 5.0 ± 2.0 |

Follow-up questionnaire waves are named according to their week and year of release (for instance, “20/26” for the follow-up questionnaire proposed to participants in the 26^th^ week of 2020).

**Table S4b.** GAD-7 score, PHQ-9 score, and Kessler-6 score throughout the first year of prospective follow-up of the COHESION Study Phase 1: detailed statistics.

| **Survey** | ***N*** | **Min** | **Q1** | **Q2** | **Q3** | **Max** | **Mean ± SD** |
| --- | --- | --- | --- | --- | --- | --- | --- |
| **Anxiety symptoms (GAD-7 score)** | | | | | | | |
| 20/26 | 128 | 0 | 2.0 | 5.0 | 9.3 | 21 | 6.1 ± 5.7 |
| 20/30 | 160 | 0 | 2.0 | 5.0 | 8.3 | 21 | 5.8 ± 4.9 |
| 20/38 | 282 | 0 | 2.0 | 5.0 | 8.0 | 21 | 6.0 ± 5.4 |
| 20/52 | 326 | 0 | 1.0 | 4.0 | 9.0 | 21 | 6.1 ± 5.8 |
| 21/15 | 332 | 0 | 2.0 | 5.0 | 10.0 | 21 | 6.4 ± 5.8 |
| 21/19 | 322 | 0 | 1.0 | 5.0 | 9.0 | 21 | 6.1 ± 5.7 |
| 21/23 | 315 | 0 | 1.0 | 4.0 | 8.0 | 21 | 5.7 ± 6.0 |
| 21/27 | 288 | 0 | 1.0 | 4.0 | 7.0 | 21 | 5.3 ± 5.4 |
| **Depression symptoms (PHQ-9 score)** | | | | | | | |
| 20/26 | 128 | 0 | 2.0 | 5.0 | 9.0 | 26 | 6.6 ± 5.8 |
| 20/30 | 160 | 0 | 3.0 | 6.0 | 9.3 | 25 | 7.0 ± 5.7 |
| 20/38 | 282 | 0 | 2.0 | 5.0 | 9.0 | 27 | 6.6 ± 6.0 |
| 20/52 | 326 | 0 | 2.0 | 5.0 | 9.0 | 27 | 6.6 ± 6.1 |
| 21/15 | 332 | 0 | 2.0 | 5.0 | 10.0 | 27 | 7.0 ± 6.4 |
| 21/19 | 322 | 0 | 2.0 | 5.0 | 9.0 | 27 | 6.8 ± 6.4 |
| 21/23 | 315 | 0 | 2.0 | 4.0 | 9.0 | 27 | 6.4 ± 6.5 |
| 21/27 | 288 | 0 | 2.0 | 4.0 | 9.0 | 27 | 6.2 ± 6.4 |
| **Psychological distress (Kessler-6 score)** | | | | | | | |
| 20/32 | 227 | 0 | 2.0 | 5.0 | 9.0 | 24 | 6.5 ± 5.6 |
| 20/34 | 294 | 0 | 2.0 | 5.0 | 9.0 | 24 | 6.4 ± 5.5 |
| 20/36 | 284 | 0 | 2.0 | 5.0 | 9.0 | 24 | 6.3 ± 5.6 |
| 20/38 | 282 | 0 | 1.0 | 4.0 | 9.0 | 24 | 6.0 ± 5.8 |
| 20/40 | 290 | 0 | 2.0 | 4.0 | 9.0 | 24 | 6.2 ± 5.7 |
| 20/44 | 348 | 0 | 2.0 | 5.0 | 10.0 | 24 | 6.4 ± 5.4 |
| 20/48 | 363 | 0 | 2.0 | 5.0 | 9.0 | 24 | 6.5 ± 5.6 |
| 20/52 | 326 | 0 | 1.0 | 4.0 | 9.0 | 24 | 5.8 ± 5.7 |
| 21/03 | 334 | 0 | 2.0 | 5.0 | 9.0 | 24 | 6.0 ± 5.3 |
| 21/07 | 362 | 0 | 2.0 | 5.0 | 9.0 | 24 | 6.4 ± 5.3 |
| 21/11 | 328 | 0 | 2.0 | 4.0 | 9.0 | 24 | 6.0 ± 5.6 |

Follow-up questionnaire waves are named according to their week and year of release (for instance, “20/26” for the follow-up questionnaire proposed to participants in the 26^th^ week of 2020).

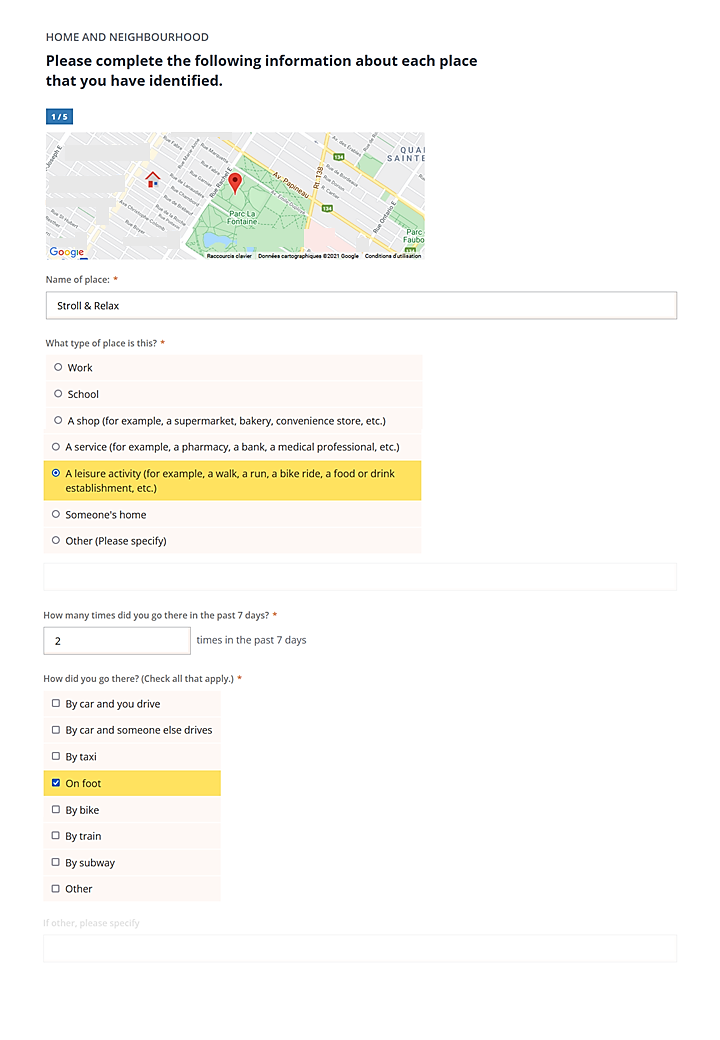

**Figure S1a.** Collecting data on each activity place with VERITAS-Social.

There is the example of a fictional participant.

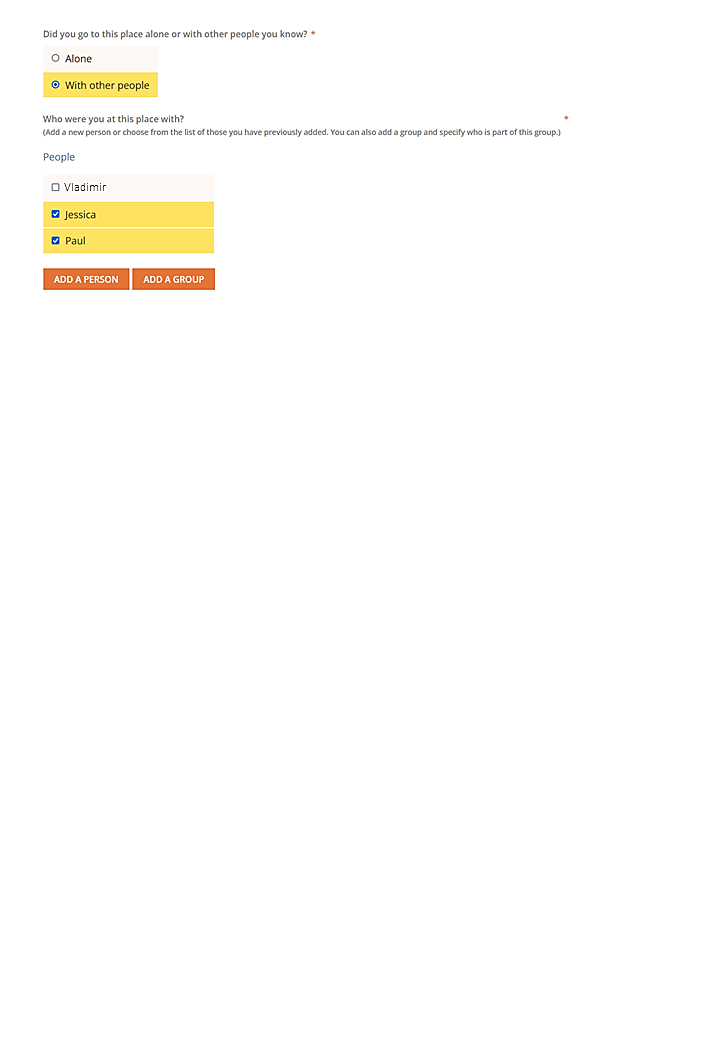

**Figure S1b.** Identifying people related to each activity place with VERITAS-Social.

There is the example of a fictional participant.

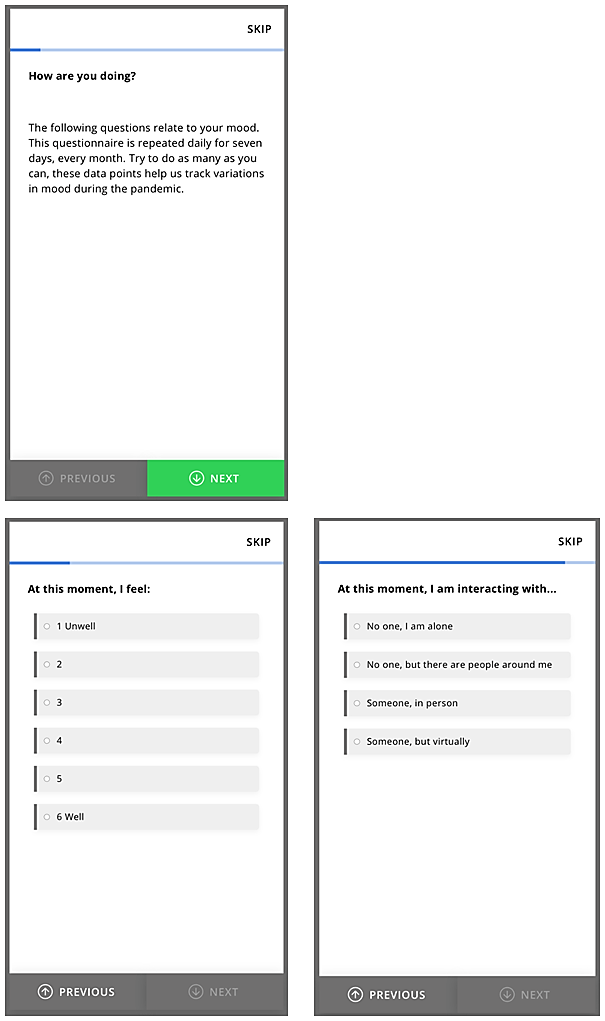

**Figure S2.** Examples of Ecological Momentary Assessment (EMA) questionnaires proposed by Ethica Data mobile application.
